# A genetically modulated Toll-like-receptor-tolerant phenotype in peripheral blood cells of children with multisystem inflammatory syndrome

**DOI:** 10.1101/2024.02.02.24301686

**Authors:** Rehan Khan, Weizhen Ji, Jeisac Guzman-Rivera, Abhilasha Madhvi, Tracy Andrews, Benjamin Richlin, Christian Suarez, Sunanda Gaur, William Cuddy, Aalok R. Singh, Hulya Bukulmez, David Kaelber, Yukiko Kimura, Usha Ganapathi, Ioannis E. Michailidis, Rahul Ukey, Sandra Moroso-Fela, John K. Kuster, Myriam Casseus, Jason Roy, Lawrence C. Kleinman, Daniel B. Horton, Saquib A. Lakhani, Maria Laura Gennaro

**Affiliations:** Public Health Research Institute, Rutgers New Jersey Medical School, Rutgers Biomedical and Health Sciences, Newark, NJ; Pediatric Genomics Discovery Program, Department of Pediatrics, Yale University School of Medicine, New Haven, CT 06510; Department of Biostatistics and Epidemiology, Rutgers School of Public Health, Piscataway, NJ; Pediatric Clinical Research Center, and Clinical Research Center, Rutgers Robert Wood Johnson Medical School, New Brunswick, NJ; Department of Pediatrics, Clinical Research Center, Rutgers Robert Wood Johnson Medical School, New Brunswick, NJ; Maria Fareri Children’s Hospital, New York Medical College, Valhalla, NY; New York Medical College, Valhalla, NY; Department of Pediatrics, Division of Rheumatology, MetroHealth System; Center for Clinical Informatics Research and Education, MetroHealth System; Department of Internal Medicine, Pediatrics, and Population and Quantitative Health Sciences, Case Western Reserve University, Cleveland OH; Hackensack University Medical Center, Hackensack Meridian School of Medicine, Nutley, NJ; Department of Pediatrics, Rutgers Robert Wood Johnson Medical School, New Brunswick, NJ; Department of Global Urban Health, Rutgers School of Public Health, Piscataway, NJ; Rutgers Center for Pharmacoepidemiology and Treatment Science, Institute for Health, Health Care Policy and Aging Research, New Brunswick, NJ; Department of Medicine, Rutgers New Jersey Medical School, Rutgers Biomedical and Health Sciences, Newark, NJ

## Abstract

Dysregulated innate immune responses contribute to multisystem inflammatory syndrome in children (MIS-C), characterized by gastrointestinal, mucocutaneous, and/or cardiovascular injury occurring weeks after SARS-CoV-2 exposure. To investigate innate immune functions in MIS-C, we stimulated *ex vivo* peripheral blood cells from MIS-C patients with agonists of Toll-like receptors (TLR), key innate immune response initiators. We found severely dampened cytokine responses and elevated gene expression of negative regulators of TLR signaling. Increased plasma levels of zonulin, a gut leakage marker, were also detected. These effects were also observed in children enrolled months after MIS-C recovery. Moreover, cells from MIS-C children carrying rare genetic variants of lysosomal trafficking regulator (*LYST*) were less refractory to TLR stimulation and exhibited lysosomal and mitochondrial abnormalities with altered energy metabolism. Our results strongly suggest that MIS-C hyperinflammation and/or excessive or prolonged stimulation with gut-originated TLR ligands drive immune cells to a lasting refractory state. TLR hyporesponsiveness is likely beneficial, as suggested by excess lymphopenia among rare *LYST* variant carriers. Our findings point to cellular mechanisms underlying TLR hyporesponsiveness; identify genetic determinants that may explain the MIS-C clinical spectrum; suggest potential associations between innate refractory states and long COVID; and highlight the need to monitor long-term consequences of MIS-C.

---

Infection with severe acute respiratory syndrome coronavirus 2 (SARS-CoV-2) is often asymptomatic or mild in children (1–4). However, pediatric SARS-CoV-2 infection may have severe consequences. The most dramatic one is a rare syndrome with multiorgan involvement that manifests several weeks after infection with SARS-CoV-2. This clinical entity, named multisystem inflammatory syndrome in children (MIS-C), was first reported in the UK early in the pandemic (5) and subsequently defined by the US Center for Disease Control and Prevention and the World Health Organization (6, 7). Symptoms of MIS-C include fever, rash, gastrointestinal symptoms, coagulopathy, myocardial dysfunction, and/or shock (4, 8). Underlying the clinical symptoms of MIS-C are multilineage immune activation and tissue inflammation, including increased gastrointestinal permeability, elevated circulating markers of microangiopathy, and release of troponin and natriuretic peptides, indicative of cardiac inflammation (4, 8). Among the immunological abnormalities reported for MIS-C, lymphopenia is most characteristic, as it separates MIS-C from the phenotypically similar Kawasaki disease and correlates with the degree of MIS-C severity (4, 8). Still, the mechanisms of MIS-C pathogenesis remain poorly understood (9). Unraveling the pathophysiology of MIS-C requires a better understanding of the underlying immune dysfunctions.

Multiple immunological studies have highlighted the contribution of innate immune cells to the systemic inflammation that is characteristic of MIS-C (4, 10). For example, severe myocarditis in MIS-C correlates with specific inflammatory gene signatures in monocytes and dendritic cells (11). In addition, studies on the genetic risk factors for MIS-C overwhelmingly point to innate immune responses as key to its pathogenesis (12–14). Our current understanding of MIS-C immunology derives from the comprehensive immunological phenotyping of peripheral blood leucocytes in their basal (i.e., non-perturbed experimentally) state by transcriptomics and multiparameter flow cytometry (15–22). Except for a few case reports (12, 13), however, studies probing innate immune cell functions in MIS-C are lacking.

To characterize innate immune responses in MIS-C, we investigated the responses triggered by engagement of Toll-like receptors (TLR). TLR are membrane proteins that bind molecules produced by pathogens or released by damaged cells and initiate downstream signaling pathways leading to the increased production of antimicrobial mediators, inflammatory cytokines, chemokines, interferons (IFNs), and co-stimulatory molecules, with subsequent development of adaptive immune responses (23–26). We report that MIS-C is associated with dampened cytokine responses to TLR stimulation and increased expression of negative regulators of TLR signaling. These phenotypes resemble TLR tolerance, a phenomenon characterized by the inability of innate immune cells to respond to further challenge when subjected to excessive or prolonged TLR stimulation. We also find that children with MIS-C exhibit elevated plasma levels of zonulin, a marker of gut permeability, as previously observed (27). Gut leakage may favor microbial translocation from the gut to the bloodstream, with consequent excessive and/or prolonged exposure of circulating blood cells to TLR ligands. We also report that among the least TLR-refractory cells are those from children carrying rare variants in the lysosomal trafficking regulator (*LYST*) gene locus. LYST, a multidomain protein implicated in various aspects of vesicular trafficking, affects lysosome morphology and function (28, 29). Pathogenic *LYST* variants are linked with Chediak-Higashi syndrome (CHS), a rare autosomal recessive disorder characterized by oculocutaneous albinism, bleeding diathesis, and congenital immunodeficiency (30, 31). Moreover, CHS and some *LYST* variants are associated with risk for hemophagocytic lymphohistiocytosis (HLH), a severe (often fatal) systemic disorder characterized by hyperinflammation (32, 33). Increased cytokine production (relative to the rest of the MIS-C group) in rare *LYST* variant carriers was associated with abnormalities in late endosome/lysosome compartment and mitochondria and defective mitochondrial energy metabolism. Moreover, rare *LYST* variant carriers tended to exhibit unfavorable clinical laboratory indicators of inflammation, supporting the possibility that TLR hyporesponsiveness may be protective during MIS-C. Our work reveals a novel aspect of the innate immune response during MIS-C and points to causative mechanisms and functional requirements. It also identifies genetic determinants of the expression of this refractory state, which may help explain the severity spectrum of the clinical manifestations of MIS-C.

## Results

### Dampened cytokine response to LPS stimulation of peripheral blood mononuclear cells from children with MIS-C

To assess whether innate immune functions are altered by MIS-C, we subjected peripheral blood mononuclear cells (PBMC) from 17 children with MIS-C to stimulation with *E. coli* lipopolysaccharide (LPS), which is a potent TLR4 agonist (34). For these children, peripheral blood had been collected within 30 days after hospital admission. Compared with PBMC from healthy controls of pediatric age, PBMC from children with MIS-C produced much lower levels of key proinflammatory cytokines, such as TNF-α, IL-6, and IL-1β (e.g., >200-fold median decrease for TNFα), in response to LPS stimulation (**Fig. 1A** for heatmap and **Fig. 1B** for boxplots). Similar results were obtained with IL-12p40 (**Fig. 1A**), which is another pro-inflammatory cytokine, and the anti-inflammatory cytokine IL-10 (Fig. S1A), which is also induced in LPS-stimulated macrophages (35, 36). The diminished cytokine response to LPS stimulation was also observed by qPCR analysis of the expression levels of corresponding proinflammatory cytokine genes (**Fig. 1C** for heatmap and **Fig. 1D** for boxplots) and for IL-10 (Fig. S1B), consistent with altered TLR signaling (25, 26). These data show that peripheral blood cells from children with MIS-C are hyporesponsive to TLR4 stimulation.

**Figure 1.**
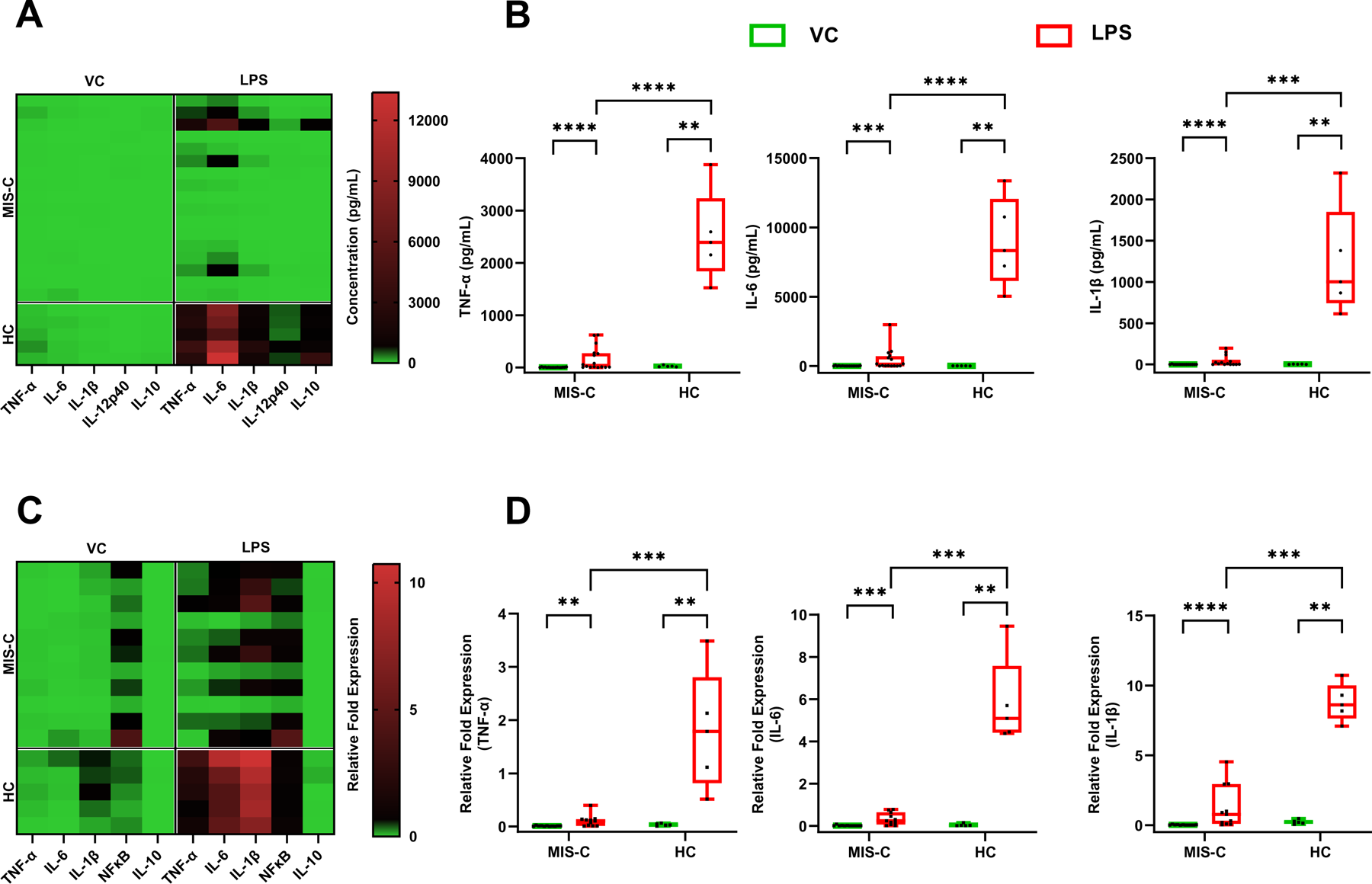
Cytokine production by peripheral blood mononuclear cells (PBMC) stimulated with LPS. PBMC collected from MIS-C patients within 30 days of hospital admission (n = 17) and from healthy controls (HC) (n = 5) were thawed and incubated with lipopolysaccharide (LPS, 50 ng/ml) or endotoxin-free water (vehicle control); cells for RNA extraction were collected after 4 hrs of treatment; cytokine release in the culture supernatant was measured after 24 hrs of treatment. All data were generated in technical duplicates. **A-B**. <u>Cytokine release</u>. Levels in culture supernatant was measured for several cytokines, as indicated, utilizing a Luminex® xMAP® platform and shown as heatmap, with cytokine levels in pg/ml (columns) across samples (rows), and gradient colors from green (low) to red (high) (panel A). The results for three pro-inflammatory cytokines are also shown as box plots (panel B); additional cytokine data are shown in **Fig. S1**. **C-D.** <u>mRNA quantification</u>. Expression levels of selected cytokine genes were quantified by qPCR and normalized to the control gene GAPDH. Results are shown as heatmap, with normalized gene (columns) expression levels across samples (rows) (panel C). mRNA levels for the corresponding inflammatory cytokines (panel B) are shown as boxplots in panel D. Box plots in panels B and D show median value, 25^th^ and 75^th^ percentile, and min/max values (whiskers); black dots represent the mean of technical duplicates per study donor. *, *p* < 0.05; **, *p* < 0.001, ***, *p* = 0.0001, ****, p <0.0001, ns, not significant. Mann-Whitney U test was used for group comparisons. MIS-C, MIS-C patients within 30 days of hospital admission; HC, healthy controls.

### Reduced TLR responses are associated with increased gene expression of negative regulators of TLR signaling and plasma indicators of abnormal gut permeability and hyperinflammation

The observed TLR hyporesponsiveness of MIS-C PBMC recalls TLR tolerance, a long-recognized phenomenon observed in severe conditions, such as sepsis, in which prior exposure of monocytes/macrophages to microbial products results in dampened TLR responses to re-stimulation of the same or a different TLR (37, 38). Since this type of immune tolerance has been best studied, although is not limited to, LPS (endotoxin), it is often referred to as endotoxin tolerance (38). Decreased cytokine production in endotoxin-tolerant cells is associated with increased expression of negative regulators of TLR signaling, including Toll-interacting protein (TOLLIP), suppressor of cytokine signaling (SOCS) 1, IL-1R-associated kinase-M (IRAK-M), sterile alpha and armadillo motif-containing molecule 1 (SARM1), and SH2 domain-containing inositol 5’-phosphatase 1 (SHIP-1) (39). When we used qPCR to measure the abundance of the corresponding transcripts in PBMC from MIS-C patients, we observed that the baseline (i.e., unstimulated) mRNA levels of SOCS1, TOLLIP, and SHIP1 were higher in the MIS-C patient cells relative to healthy controls (the comparison for SARM1 was close to significance, *p* = 0.06) (**Fig. 2A** for heatmap and **Fig. 2B** for boxplots). Increased expression of negative regulators of TLR signaling is consistent with the dampened response of MIS-C cells to TLR4 stimulation by LPS.

**Figure 2.**
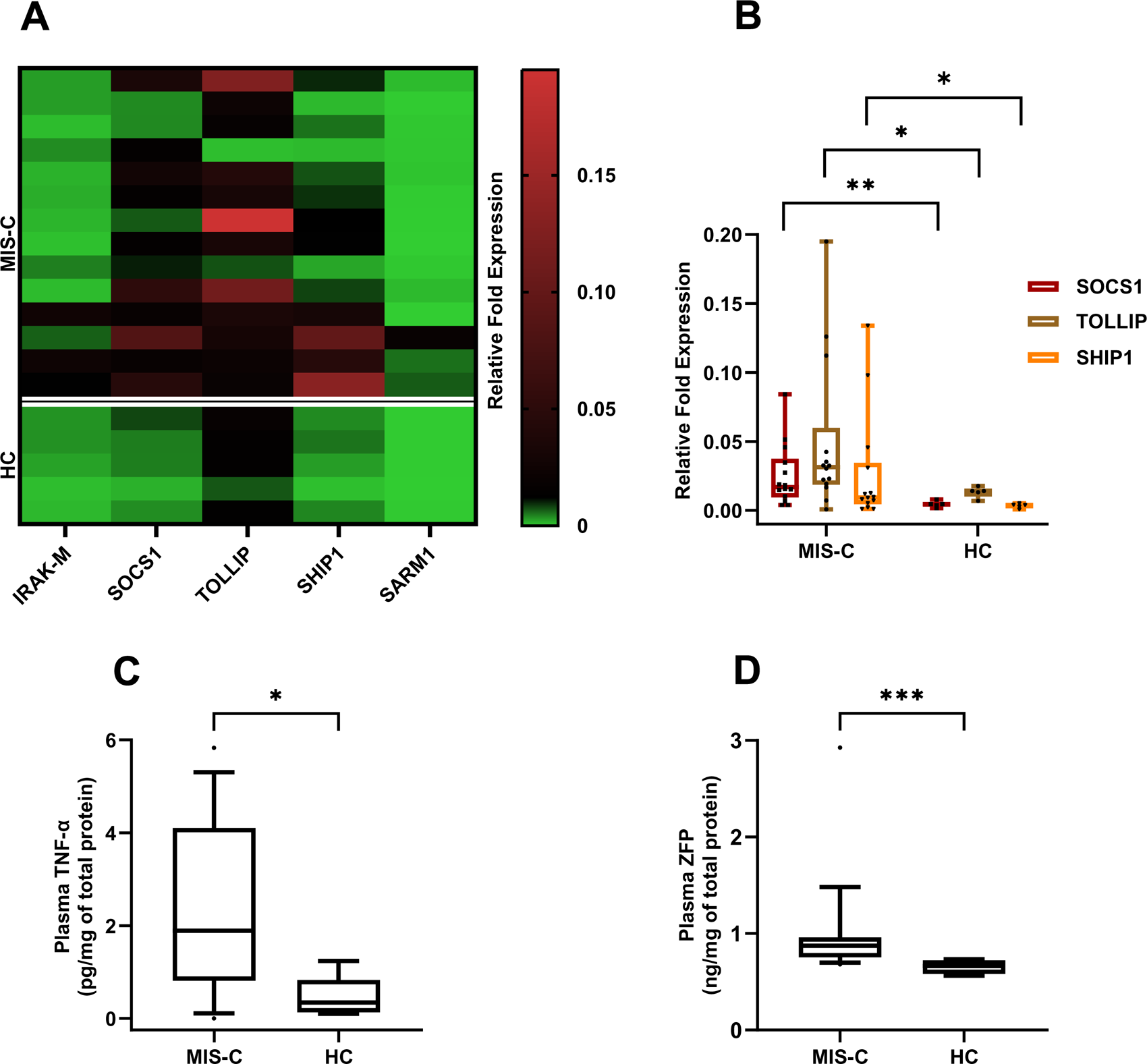
Mechanisms potentially responsible for TLR hypo-responsiveness. PBMC (panels A-B) and plasma (panels C-D) were obtained from peripheral blood of MIS-C patients (n = 17) and healthy controls (HC) (n = 5), as in Fig. 1. **A-B.** <u>Expression</u> <u>levels of genes encoding negative regulators of TLR signaling</u>. RNA for qPCR of selected TLR signaling regulator genes was obtained from untreated PBMC collected from MIS-C patients and healthy controls (HC), as in Fig. 1. Results are shown as heatmap (Panel A), with normalized gene (columns) expression levels across samples (rows). For selected genes, results are also shown as box plots (panel B), as in Fig. 1. **C.** <u>Plasma cytokine levels</u>. The levels of major cytokines were measured in technical duplicates by utilizing a Luminex® xMAP® platform and results were normalized to total protein in the sample. **D.** <u>Plasma zonulin levels</u>. The levels of zonulin were measured in technical duplicates by using a commercial zonulin family peptide assay kit and results were normalized to total protein in the sample, as in *Methods*. Box plots show median value, 25^th^ and 75^th^ percentile, and 10-90 percentile values (whiskers). *, *p* < 0.05; **, *p* < 0.001, ***, *p* =0.0001, ****, p <0.0001, ns, not significant (Mann-Whitney U test).

We reasoned that the refractory state of the innate immune cells in MIS-C patients could be explained by at least two, non-exclusive scenarios. One relates to the excessive production of proinflammatory molecules, including cytokines, characteristic of MIS-C (4) (examples of cytokine levels in plasma of children with MIS-C are shown in **Fig. 2C** and **Table S2**). High cytokine levels might induce negative feedback mechanisms including the upregulation of negative regulators to diminish further cytokine production. This type of mechanism has been reported for SOCS1 (40). A second scenario is suggested by the finding that MIS-C is accompanied by loss of gut mucosal barrier, as indicated by the release into the bloodstream of zonulin, a biomarker of intestinal permeability (27). Consistent with this report, we found that the plasma levels of zonulin were higher in children with MIS-C, relative to healthy controls (**Fig. 2D**). MIS-C-induced loss of gut mucosal barrier supports the possibility that microbial products released from the gut into the bloodstream induce excessive and/or prolonged stimulation of circulating innate immune cells and consequent TLR hyporesponsiveness. Lastly, our data do not support abnormal TLR surface expression as driver of reduced TLR responsiveness, since the abundance of TLR4 on the surface of PBMC from children with MIS-C was indistinguishable from healthy controls (**Fig. S2**).

### Reduced TLR responses and associated markers are also observed with blood samples obtained >30 days after hospital admission for MIS-C

We had available for analysis PBMC and plasma samples obtained from a group of 16 children recovered from MIS-C, for whom peripheral blood was collected greater than 30 days after hospital admission [median (IQR) = 202 (81.5, 444) days]. We were surprised to observe reduced cytokine production in response to LPS stimulation also in these children, relative to healthy controls (**Fig. S3A**). In addition, transcript levels of SOCS1, TOLLIP, SARM1, and SHIP1 (**Fig. S3B**) and plasma levels of zonulin (**Fig. S3C**) were higher in this group than in healthy controls. Together, these results suggest that TLR hyporesponsiveness and gut barrier leakage associated with MIS-C are lasting.

### The cytokine response to TLR stimulation of peripheral blood mononuclear cells from children with MIS-C shows a positive skew

We compared the distribution of the cytokine data obtained in response to LPS stimulation in MIS-C and healthy control groups. We observed that the MIS-C data distribution was highly skewed, with an abundance of very low values and a tail of higher values (see the IL-6 example in **Fig. 3A**, left panel), unlike the normal distribution observed with the healthy control data (**Fig. 3A**, right panel). The distribution differences are likely non-random, since the sample size with MIS-C in the study (n = 33) exceeds that of the healthy control group (n = 5), and samples of small sizes are less likely to be normally distributed (41). Moreover, the positive skew of cytokine data was reflected by greater dispersion of the data (coefficients of variation, CV >1) in the MIS-C group than in the healthy control group (CV <1) (**Fig. 3A**, insets). Furthermore, the two data distributions were essentially non-overlapping: with the exception of a single outlier, all MISC values were lower than the lowest healthy control (**Fig. 3A**). Thus, the two distributions were different in shape, dispersion, and magnitude.

**Figure 3.**
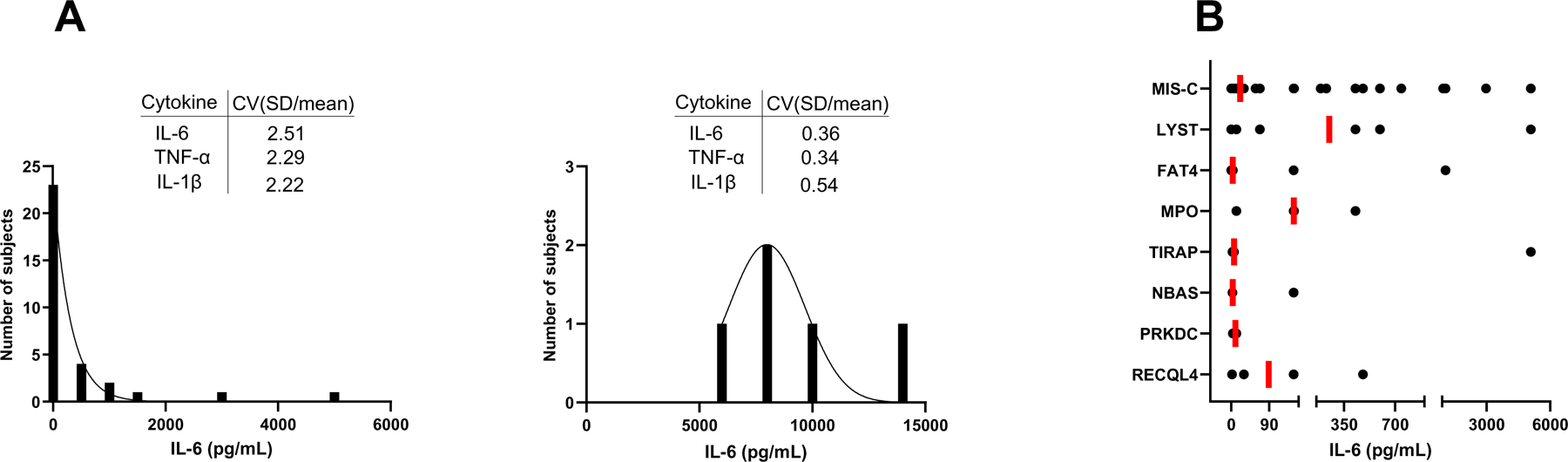
Distribution of cytokine release data and gene involvement. **A.** <u>Frequency distribution for IL-6 cytokine data:</u> IL-6 cytokine levels in culture supernatants of PBMC incubated with 50 ng/ml LPS for 24 hr were measured as in Fig. 1. The graph was obtained by using frequency distribution analysis and a Gaussian distribution model in GraphPad Prism; MIS-C (n = 33), left panel; healthy controls (HC) (n = 5), right panel. In both panels, the inset shows the coefficient of variation (CV) (SD/mean) for three proinflammatory cytokines**. B.** <u>Cytokine level and its distribution</u> <u>range in subjects with most frequently recurrent immunity-related target genes in the</u> <u>MIS-C group</u>. The figure shows the distribution of data on IL-6 release from LPS-stimulated PBMC, as in Panel A, for all MIS-C subjects (top row) and by each of the seven most frequently recurrent immunity-related target genes among MIS-C subjects (subsequent rows, as indicated) from a list of 623 genes (**Table S4**). No data points are removed by the *x* axis breaks, which were introduced to improve data visualization. One black dot represents one donor; the vertical red line represents the median of the distribution. The highest value in the *LYST* and *TIRAP* rows corresponds to the same donor. *LYST*, lysosomal trafficking regulator; *FAT4*, FAT atypical cadherin 4; *MPO*, myeloperoxidase; *TIRAP*, TIR domain containing adaptor protein; *NBAS*, NBAS subunit of NRZ tethering complex; *PRKDC*, protein kinase, DNA-activated subunit; *RECQL4*, ReqQ-like helicase.

### Rare variants in *LYST* are associated with the skewed cytokine data distribution in MIS-C

We examined the variables underlying the data distribution in the MIS-C group. Differences in cytokine responses to TLR stimulation among children with MIS-C were not associated with demographic characteristics (such as sex at birth, age, race, or ethnicity) or time elapsed between MIS-C hospital admission and blood draw (≤ 30 days vs > 30 days) (**Table S3**). Thus, we next considered the genetic background of the study participants. We analyzed germline whole genome sequencing for children with MIS-C. Within a list of 623 genes related to immune function (**Table S4**), we focused on rare (minor allele frequency (MAF) <0.005) non-synonymous variants and further screened for a Combined Annotation Dependent Depletion (CADD) score of ≥20 to identify missense variants that were more likely to impact protein function (42, 43). We found that seven of the 623 genes exhibited four or more non-synonymous, rare variants in the 33 MIS-C patients (**Tables S5-6**). We then examined the distribution of cytokine data in MIS-C patients grouped based on the presence of rare variants in each of these genes. We observed that the carriers of rare non-synonymous *LYST* variants (n = 7) (**Table S6**) showed the highest median, driving the high values in the MIS-C distribution (see the IL-6 example in **Fig. 3B**). Moreover, the rare *LYST* variant data clustered in three groups (low, medium, and high) that mimicked those seen in the MIS-C patients. This result shows the *LYST* genotype contributes to the skewed distribution of the cytokine data in the MIS-C group.

### Rare non-synonymous *LYST* variants are associated with lysosomal and mitochondrial abnormalities, altered energy metabolism, and unfavorable clinical laboratory indicators

The observed skew in the cytokine response prompted investigation of cellular phenotypes associated with rare *LYST* variants. Since LYST function has been associated with lysosome morphology and function (28, 29), we tested PBMC of rare non-synonymous *LYST* variant carriers (**Table S6**) for lysosome marker. For this analysis, the similarly sized comparator group was constituted by children with MIS-C carrying common (MAF >0.005) non-synonymous *LYST* variants (**Table S7**). The two subsets of children did not differ in terms of basic demographic characteristics (except race) and treatments received during MIS-C hospitalization (**Table S8**). By using imaging flow cytometry, we found that the rare *LYST* variants in the MIS-C group were selectively associated with increased abundance of lysosomal-associated membrane protein 1 (LAMP1) in leukocytes (**Fig. 4A** for box plots and for selected images), indicative of altered morphology and/or function of late endosomes and lysosomes in these cell types (29, 44). This result is consistent with the organelle enlargement known to occur in Chediak-Higashi syndrome (29). Lysosomal functions are intricately interconnected with mitochondrial functions (45), which in turn shape cellular metabolism as well as immune cell functions (46, 47). Indeed, we found that cell types showing altered LAMP1 staining in the rare *LYST* variant group also exhibited decreased labeling with MitoTracker Red (**Fig. 4B**), a dye for active mitochondria labeling. Mitochondrial damage in the rare *LYST* variant carriers was further indicated by reduced mitochondrial metabolism (oxidative phosphorylation) accompanied by increased non-mitochondrial metabolism (glycolysis), as measured by Seahorse XF technology (**Fig. 4CD**). Collectively, the results above show that the occurrence of rare *LYST* variants is associated with altered TLR-mediated immune responses, lysosomal and mitochondrial abnormalities, and altered energy metabolism.

**Figure 4.**
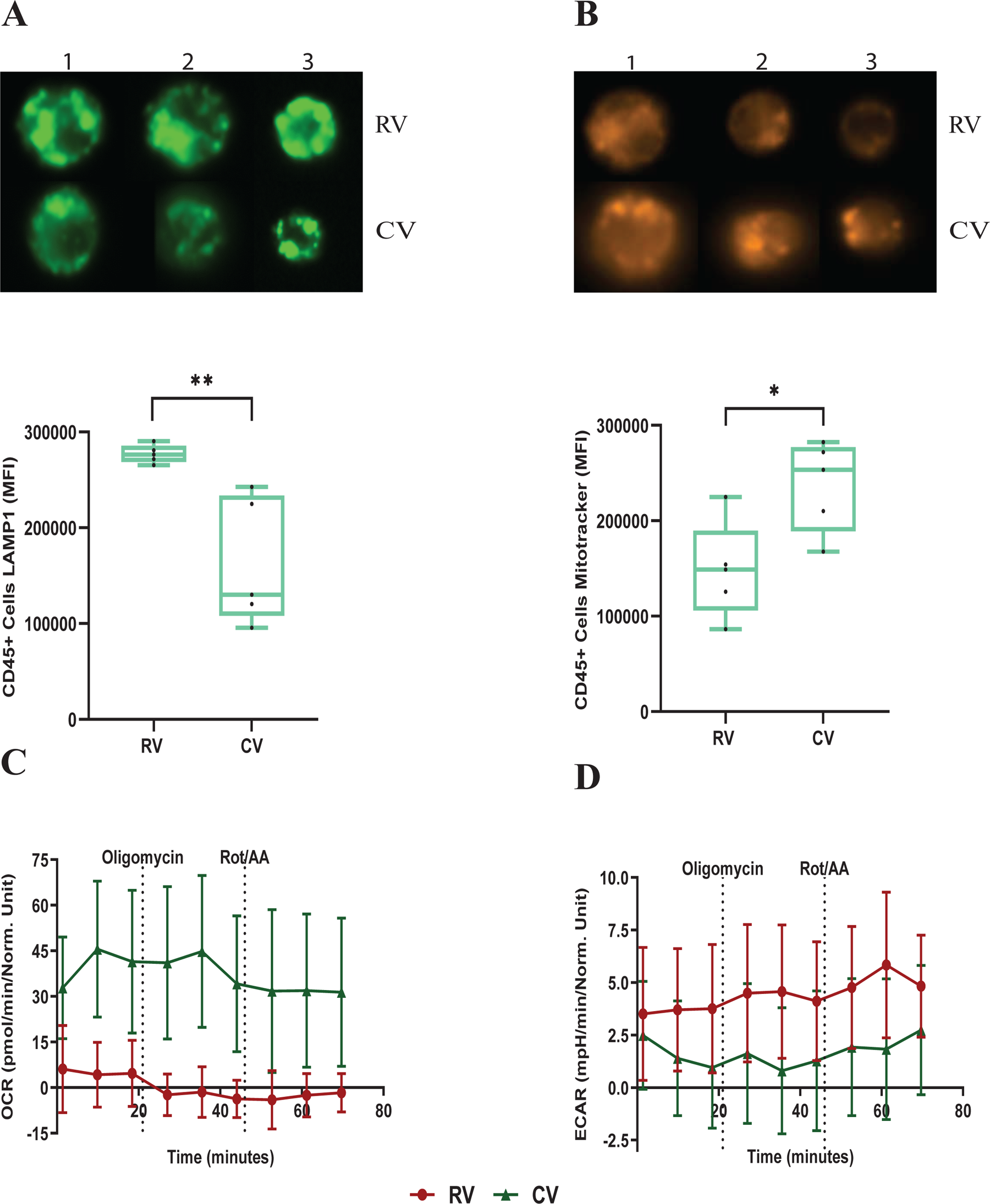
Analysis of lysosomal, mitochondrial, and metabolic markers associated with rare *LYST* variants. Comparisons in this figure included MIS-C subjects carrying rare *LYST* variants [minor allele frequency (MAF) <0.005] vs. MIS-C carriers of common *LYST* variants (MAF >0.005). RV, rare variant carriers (n = 5); CV, common variant carriers (n = 5 in panels AB and 4 in panels CD). **A**. <u>LAMP1 abundance</u> in CD45^+^ cells. Data were obtained by Imaging Flow Cytometry. The figure shows representative images of three MIS-C subjects (upper panel) and box plots with median value, 25^th^ and 75^th^ percentile, and min/max values (whiskers) (lower panel). The relative frequency of CD45+ cells showed no significant differences between the two groups in the comparison. **B.** MitoTracker intensity in CD45^+^ cells. Data were obtained by Imaging Flow Cytometry and expressed as in Fig. 4A A. *, *p* < 0.05; **, *p* < 0.001, ***, *p* =0.0001, ****, p <0.0001, ns, not significant, using Mann-Whitney U test. **C-D.** <u>Metabolic profile of PBMCs</u>. Seahorse assay results for (C) oxygen consumption rates (OCR) and (D) extracellular acidification rates (ECAR) at baseline and in response to mitochondrial inhibitors. Values are presented as mean +/-SEM. The unusual OCR and ECAR profiles result from person-to-person variability and data pooling – representative profiles obtained with single study subjects are shown in **Fig. S4**.

We next asked whether the dysfunctions observed at a cellular level in the children carrying the rare *LYST* variants were reflected in the clinical course of MIS-C. For this assessment, we analyzed the results of laboratory tests having most relevance to MIS-C, such as markers of inflammation and white blood cell counts (48). We observed numerical differences for key markers of disease severity between the rare and common *LYST* variant groups. White blood cell counts tended to be lower in the rare *LYST* variant group than in the common *LYST* variant counterpart. Neutrophil counts were only slightly reduced (<2-fold), but lymphopenia, which is a key finding of MIS-C (4, 48), was considerably more pronounced in the rare *LYST* variant group than in the common *LYST* variant group (p < 0.05). The resulting median neutrophil-to-lymphocyte ratio, which is a key marker of disease severity in many pathologies (49), was 4-fold higher in rare than common *LYST* variant carriers (**Table S9**). Despite the small size of the sample tested (n = 13 total across groups), the observed differences suggest that the rare *LYST* variants are associated with more severe clinical presentations.

## Discussion

We report that MIS-C, the multi-system hyperinflammatory syndrome that may develop in children following exposure to SARS-CoV-2, is associated with a refractory state of the innate immune cells in the bloodstream that is evidenced as inability to respond to TLR stimulation ex vivo. Increased gut permeability, as indicated by elevated plasma levels of zonulin in patients with MIS-C in this study and previous work (27), may result in microbial translocation, leading to excessive and/or prolonged exposure of circulating immune cells to TLR ligands of microbial origin and their consequent TLR tolerization. The detection of SARS-CoV-2 antigen, perhaps of intestinal origin (27), in the bloodstream of MIS-C patients supports this scenario, particularly since the SARS-CoV-2 Spike glycoprotein can act as TLR ligand (50–54). Moreover, the activation of negative feedback responses induced by high levels of proinflammatory mediators in the bloodstream likely concurs to render innate immune cells of children with MIS-C unable to respond to further challenge. We also show that, among cells from children with MIS-C, the least refractory to TLR stimulation are those carrying non-synonymous, rare variants in *LYST*. Variants in *LYST* and other genes associated hemophagocytic lymphohistiocytosis (33) have been previously associated with MIS-C risk (55). By pointing to altered morphology and/or function of mitochondria and late endosomal/ lysosomal compartments in *LYST* variant carriers, our study also identifies cellular mechanisms that may be required to establish or maintain a refractory state of innate immune cells to TLR challenge.

The finding that MIS-C is associated with dampened ability of innate immune cells to respond to stimulus contributes new understanding of the “post-COVID” immunological landscape. For example, it is postulated that TLR tolerance helps limit immune hyperactivation and uncontrolled inflammation, as mentioned above, but it is also associated with unfavorable outcomes, such as increased risk of secondary infections or chronic inflammatory states (37, 38). Thus, our results raise the question whether the increased incidence of infections observed in the post-COVID era could be attributed to increased susceptibility rather than to the “immunity debt” from insufficient immune stimulation due to non-pharmaceutical interventions used to limit the spread of COVID-19, such as masking or social isolation (56, 57). Since MIS-C is rare among the manifestations of pediatric SARS-CoV-2 infection, addressing this question requires investigating whether other forms of pediatric COVID-19 also lead to diminished TLR responses. Moreover, our data could help explain mechanisms of pathogenesis of post-acute sequelae of SARS-CoV-2 infection (PASC, also known as long COVID), which have affected 200 million people worldwide (58). We were surprised to observe that PBMC obtained several months after recovery from MIS-C exhibited similar levels of TLR hyporesponsiveness as those obtained within few weeks after hospitalization for MIS-C. Our study was not designed to monitor long-term consequences of MIS-C, so we cannot exclude alternative, albeit unlikely, explanations (e.g., that children enrolled with MIS-C history later had unrelated conditions also associated with TLR hyporesponsiveness). It is however tempting to speculate that lasting hyporesponsiveness to TLR stimulation may result from prolonged innate immune cell stimulation. At least two scenarios may explain prolonged stimulus. One is viral persistence, one of the mechanisms proposed as drivers of long COVID (59) since, as mentioned above, the Spike protein of SARS-CoV-2 can act as TLR ligand (50–54) and has been detected in the bloodstream of children with MIS-C (27). An alternative scenario would involve the lasting, epigenetic proinflammatory reprogramming of human immune stem cells induced by SARS-CoV-2 infection (60), since circulating innate immune cells may become tolerant in response to prolonged exposure to proinflammatory molecules. Thus, whether direct or indirect, relationships may exist between lasting immune stimulation, innate immune tolerance, and various manifestations of long COVID.

The potential involvement of *LYST* supports the possibility that MIS-C, at least in some cases, may be associated with lysosomal defects. Lysosomal dysfunctions underlie multiple pathological conditions, including lysosomal storage diseases, which also exhibit organelle dysfunction, mitochondrial damage, and neuroinflammation, and autoimmune diseases (61, 62). Thus, lysosome targeting drugs developed for these pathologies (62) might be considered in the management of some cases of MIS-C. Mechanistic hypotheses linking *LYST* defects and inflammation and related drug effects will be best tested with isogenic mutant immune cell lines.

In conclusion, our data identify TLR hyporesponsiveness as a novel immunological mechanism in MIS-C, where it may have beneficial or harmful consequences. On one hand, reduced responses to TLR stimulation may contribute to prevent runaway inflammation and protect against worse clinical outcomes. Indeed, the potential association between reduced TLR activation and unfavorable markers of inflammation observed in children with rare *LYST* variants supports a protective role of TLR hyporesponsiveness. Moreover, since MIS-C presents with varying degrees of clinical severity, child-to-child differential ability to express this refractory state and the underlying genetic determinants may help explain the severity spectrum of MIS-C manifestations. On the other hand, reduced ability to respond to TLR ligands, which may be prolonged, may result in increased post-COVID susceptibility to various infections and, potentially, contribute to long COVID. Our findings warrant further investigations to validate associations between genotypes and molecular and clinical phenotypes and to determine whether other clinical manifestations of SARS-CoV-2 infection are also associated with TLR hyporesponsiveness. Our work also highlights the importance of longitudinal studies to monitor the health of children who have experienced MIS-C.

## Data Availability

All data produced in the present study are available upon reasonable request to the authors

## Acknowledgments

We wish to thank Karl Drlica and George Yap for critical reading of the manuscript; Rozina Aamir, Lisa Cerracchio, Wendy Dalton, Justine Griswold, Monica Konstantino, and Emanuel Lerner for their assistance with patient referral to the study, recruitment, enrollment, collection of data and biospecimens, and regulatory documentation; Mary Ellen Riordan for her insights and operational expertise in coordinating clinical sites and recruitment; the Yale Center for Genome Analysis for DNA sequencing; Charles Hevi for coordinating biospecimen collection and shipping to biorepository with consortium members; and Beth Dworetzky and Steph Lomangino from Family Voices for their assistance and guidance with recruitment materials, surveys, and other participant-facing documents. We are indebted to the MIS-C patients and their families for consenting to participate to the study. This work was funded in part by NICHD HD105619, NIAID R01AI158911, and NCATS UL1TR003017.

## Disclosur

SAL is part owner of Victory Genomics and Qiyas Higher Health, two startup companies unrelated to this work.

## Author contributions

Design of experimental plan (RK, WJ, SAL, MLG); execution of experimental work (RK, JG-R, AM, UG, IEM, RU, WJ, JKK); participant recruitment, data collection, and biospecimen collection (BR, CS, SG, WC, ARS, HB, DK, YK); project management (SM-F); data management (MC, DH); statistical analysis (TA, JR); interpretation of results (all authors); manuscript writing (RK, SAL, MLG); critical review of manuscript (all authors); consortium leadership (MLG, DH, LCK).

## Materials and Methods

### Study Design and Population

COVID-19 Network of Networks Expanding Clinical and Translational approaches to Predict Severe Illness in Children (CONNECT to Predict SIck Children) is a multicenter prospective case-control study designed to predict children at greatest risk of severe consequences from SARS-CoV-2 infection. Eligibility criteria included: (1) confirmed diagnoses of SARS-CoV-2 infection or MIS-C; (2) age ≤ 21 years at time of enrollment; and (3) not pregnant either during the qualifying illness or at the time of enrollment. Children and youth with MIS-C were classified in accordance with the 2020 U.S. Centers for Disease Control criteria (63): (1) positivity for current or recent SARS-CoV-2 infection by RT-PCR, serology, or antigen test, or COVID-19 exposure within 4 weeks prior to onset of symptoms; (2) a combination of the following criteria: (a) fever; (b) laboratory evidence of inflammation (e.g., elevated CRP, D-dimer, IL-6); (c) evidence of clinically severe illness requiring hospitalization with dysfunction or disorders affecting at least 2 organs: cardiac, renal, respiratory, hematologic, gastrointestinal, dermatologic, or neurological; and (3) no alternative plausible diagnosis. Healthy controls were well children (3 males and 2 females) with no pre-existing chronic inflammatory illness, without an acute COVID-19 illness in the past month, serologically negative for SARS-CoV-2 infection (anti-Nucleocapsid antibodies), and no acute illness at the time of study blood sample collection.

### Study Activities and Data

Participants were enrolled between June 23, 2021, and November 24, 2022. Potential participants were identified through searches of electronic health records and recruited through direct contact in clinical settings or using flyers, phone calls, or emails. Following consent, parental permission, and/or assent as appropriate, participants with MIS-C or their parents or guardians completed a questionnaire. Clinical information (medical history, treatments, laboratory results, imaging, and other diagnostic tests) was obtained through medical chart review. Peripheral blood was collected by phlebotomy and shipped to the study biorepository. Biological parents of participants with MIS-C were also invited to participate and provide a blood or cheek swab sample for genomic sequencing. Study data were collected and managed using REDCap electronic data capture tools hosted at Rutgers Robert Wood Johnson Medical School (64, 65). A summary of demographic characteristics and MIS-C-related clinical laboratory findings is presented in **Table S1**.

### Study approval

All study activities were approved by the Rutgers Institutional Review Board (Pro2020002961) and all participants provided informed consent prior to engaging in study activities.

### Whole blood processing

Peripheral blood mononuclear cells (PBMCs) were isolated by standard Ficoll density gradient centrifugation and stored in liquid nitrogen until use. Plasma was collected and stored at -80°C until use.

### Whole genome and exome sequencing and analysis

Genomic DNA was isolated from peripheral blood of MIS-C patients and sequencing was performed by Illumina next generation sequencing under a research protocol at the Yale Center for Genome Analysis (YCGA). Whole genome sequencing (WGS) was carried on 22 samples with mean coverage 35x, and whole exome sequencing (WES) was carried on 11 samples with mean coverage 65x. Whole exome was captured using IDT xGen capture kit v.2 (66, 67). Paired end sequence reads were converted to FASTQ format and were aligned to the reference the human genome (GRCh37/hg19). SNVs and indels were called using an automated GATK-based pipeline scripted by YCGA with AnnoVar annotation. We filtered in the exonic or splice region rare variants (MAF ≤ 0.005 in gnomAD) that exhibited high quality sequence reads with pass GATK Variant Score Quality Recalibration, genotype quality score GQ ≥ 30, a minimum 10 total reads, and ≥25% alternate allele ratio. We screened the results against a panel of 623 target genes related to immune function. Truncating variants (nonsense or splicing variants and indels) and missense variants with CADD≥20 in these genes were used for rare variant analysis.

### Peripheral blood mononuclear cells (PBMC) and Toll-like receptor (TLR) stimulation

Cryopreserved PBMCs were thawed and washed in pre-warmed RPMI 1640 supplemented with 2 mM L-glutamine, 10% FBS, 100 U/ml penicillin, and 100 μg/ml streptomycin (all from Corning cellgro, Manassas, VA) (complete RPMI). PBMCs were resuspended in complete RPMI and plated in cell culture plates with different cell densities for individual experiments accordingly. Plated cells were stimulated with lipopolysaccharide (LPS; Cat. No. tlrl-peklps, InvivoGen, USA). Endotoxin-free water was used as vehicle control. Culture plates were incubated for specific time at 37°C in a 5% CO_2_ humidified atmosphere accordingly.

### Quantitative Gene Expression Analysis (Q-PCR)

Thawed and resuspended PBMCs (250K/well) were plated in cell culture plates in duplicate and stimulated with LPS (50 ng/mL) and vehicle control for 4 hours. Cells were harvested and washed with PBS, and RNA was isolated using RNeasy Plus Mini Kit (Cat. No. 74134, Qiagen, USA) and stored at -80°C. The total RNA was reverse transcribed into cDNA using the QuantiTect Reverse Transcription Kit (Cat. No. 205311, Qiagen, USA). Transcript levels for genes listed in **Table S10** were measured by q-PCR using KiCqStart® SYBR® Green qPCR ReadyMix™ (Cat. No. KCQS00, Sigma-Aldrich, USA) on LightCycler® 480 II Instrument (Roche Diagnostics, USA). GAPDH was used as internal control. Primers used in q-PCR are listed in **Table S10**.

### Multiplex cytokine analysis

Thawed and resuspended PBMCs (50,000/well) were plated in cell culture plates in duplicate and stimulated with LPS (50ng/mL) or endotoxin-free water (vehicle control) for 24 hours. After incubation, culture supernatant was collected by centrifugation at 2,000 rpm for 5 minutes and stored at -80°C. Multiplex cytokine bead assays to measure TNFα, IL-6, IL-1β, IL-12p40, and IL-10 were performed with culture supernatants and patient plasma samples using MILLIPLEX® Human Cytokine/Chemokine Multiplex Assay kits (Cat. No. HCYTA-60K, MilliporeSigma, USA). Results were analyzed with a Luminex™ xMAP™ INTELLIFLEX System. Plasma cytokine measurements were normalized to total protein in the sample.

### Zonulin ELISA

Zonulin family peptides (ZFP) levels were assayed in patient plasma samples using a commercially available ELISA kit (Cat. No. 30-ZONSHU-E01, ALPCO, USA), according to manufacturer’s instructions. Zonulin levels were normalized to total protein in the sample.

### Flow cytometry

Thawed and resuspended PBMCs (500,000/well) were plated in cell culture plates and then stimulated with LPS (50ng/mL) and endotoxin-free water (vehicle control) for 24 hours. Cells were harvested, washed with FACS buffer, centrifuged, and resuspended and incubated with Human TruStain FcX™ (Cat No. 422302, BioLegend, USA) FcR blocking solution for 15 minutes. APC anti-human CD284 (TLR4) antibody (Cat. No. 312816, BioLegend, USA) was then added and incubated for 30 minutes at 4°C. Cells were washed with FACS buffer, centrifuged, resuspended in FACS buffer and LIVE/DEAD™ Fixable Blue Stain (Cat. No. L23105, Thermo Fisher Scientific Inc., USA), and incubated in the dark for 30 minutes at 4°C. Cells were washed with FACS buffer, centrifuged, resuspended in Fixation/ Permeabilization solution (Cat. No. 554714, BD Biosciences, USA), and incubated for 20 minutes at 4°C. Cells were washed with BD Perm/Wash™ buffer and resuspended in FACS buffer. Data were acquired in a BD LSRFortessa X-20 flow cytometer (BD Biosciences, San Jose, CA) and analyzed with FlowJo 10.9 software (FlowJo). BD™ Cytometer Setup & Tracking beads (CS&T, BD Biosciences) were used to calibrate the flow cytometer before each experiment. A total of 100,000–200,000 events were acquired per sample. Polystyrene compensation beads (BD Biosciences) were used to calculate spectral overlap values for each fluorochrome, according to the manufacturer’s instructions.

### Imaging flow cytometry

Thawed and resuspended PBMCs (500,000/well) were plated in cell culture plates and stimulated with LPS (50ng/mL) and endotoxin-free water (vehicle control) for 24 hours. Cells were harvested, washed with PBS, centrifuged, resuspended in FACS buffer with Human TruStain FcX™ (Cat No. 422302, BioLegend, USA) FcR blocking solution and incubated for 15 minutes. PE anti-human CD45 antibody (Cat. No. 304008, BioLegend, USA) was then added and incubated for 30 minutes at 4°C. Cells were washed with FACS buffer, centrifuged, resuspended, and incubated with BD Horizon™ Fixable Viability Stain 780 (Cat. No. 565388, BD Biosciences, USA) in the dark for 30 minutes at 4°C. Cells were washed with FACS buffer, centrifuged, resuspended in PBS with MitoTracker™ Red CMXRos staining dye (Cat No. M7512, Invitrogen™, USA), and incubated for 30 minutes at 37°C in a 5% CO_2_ humidified atmosphere. Cells were washed with PBS, centrifuged, resuspended in Fixation/ Permeabilization solution (Cat. No. 554714, BD Biosciences, USA), and incubated for 20 minutes at 4°C. Cells were washed with BD Perm/Wash™ buffer, resuspended in PBS, and incubated with FITC anti-human CD107a (LAMP-1) antibody (Cat. No. 328606, BioLegend, USA) for 30 minutes at 4°C. Data from 5,000–10,000 cells were acquired with an ImageStreamXMark II imaging flow cytometer (Amnis Corporation, Seattle, WA) using 60× magnification. Image data were analyzed by IDEAS software version 6.2 (Amnis Corporation, Seattle, WA) after applying a compensation matrix.

### Measurement of oxidative phosphorylation and glycolysis in PBMCs

Thawed and resuspended PBMCs (500,000/well) were plated in Seahorse culture plates and cultured for 24 hours at 37°C in a 5% CO_2_ humidified atmosphere. Extracellular acidification rates (ECAR) and oxygen consumption rates (OCR) were measured in XF media (nonbuffered RPMI 1640 containing 10mM glucose, 2mMLL-glutamine, and 1mM sodium pyruvate) under basal conditions and in response to 1.5μM oligomycin (ATP synthase inhibitor), and 0.5μM Rotenone and Antimycin A (Rot/AA; inhibitors of electron transport chain) by using the Seahorse XFe24 Analyzer (Agilent Technologies), according to the manufacturer’s instructions. For data normalization in each well, cells were incubated with DAPI dye after measurement of OCR and ECAR for 15 minutes at room temperature in dark. Plates were imaged using BioTek Cytation 5 Cell Imaging Multimode Reader (Agilent), and cell numbers per well were determined by using BioTek Gen5 Software for Imaging & Microscopy (Agilent).

### Statistical Analysis

Data were analyzed and visualized with GraphPad Prism version 10 and STATA 18. Categorical variables were presented as frequencies and percentages; differences across groups were tested with Chi-Square or Fisher Exact tests, as appropriate.

Continuous variables were summarized as medians with interquartile range; normality was assessed using a Kolmogorov-Smirnov test (p<0.001). As these variables were non-parametric, differences across groups were tested using a Mann-Whitney U test or a Kruskal Wallis test, as appropriate. Box plots were drawn to show the 25th percentile, the median, and the 75th percentile of the distribution. To highlight differences in the distribution of data across groups, the coefficient of variation was calculated from the mean and standard deviation of the distribution and inset in the plots. Significance was set as *p* < 0.05 for all tests. No data were excluded from the analysis.

## Supplementary figure legends

**Figure S1.**
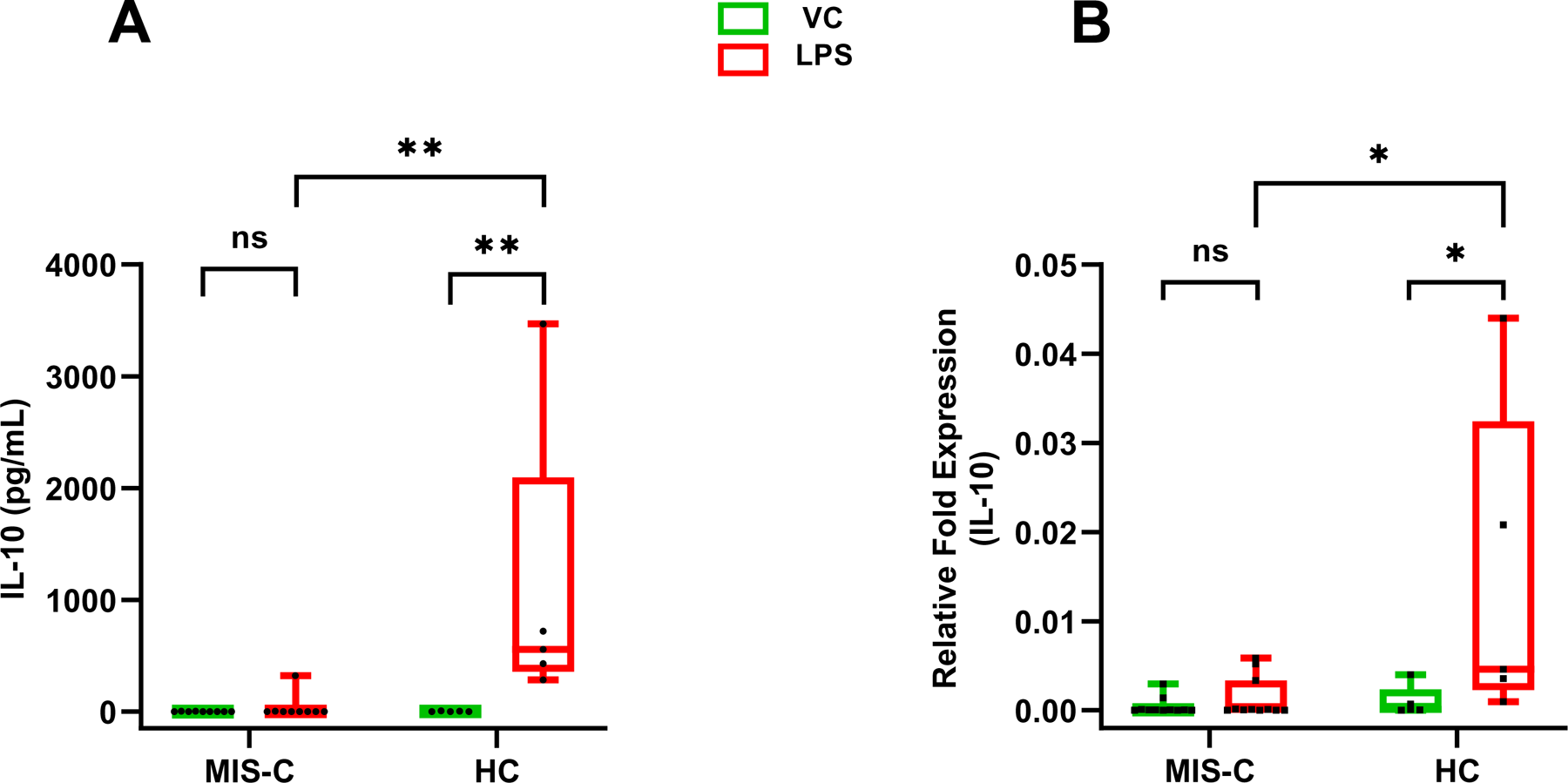
IL-10 release and mRNA measurements in peripheral blood mononuclear cells (PBMC) stimulated with LPS. PBMC collected from MIS-C patients (n = 17) and healthy controls (HC) (n = 5) were thawed and incubated with lipopolysaccharide (LPS, 50 ng/ml) or endotoxin-free water (vehicle control); A. Cytokine release assays of the culture supernatants and B. Measurements of transcript abundance were performed as in Fig. 1. *, *p* < 0.05; **, *p* < 0.001, ns, not significant. Mann-Whitney U test was used for group comparisons. MIS-C, MIS-C patients within 30 days of hospital admission; HC, healthy controls.

**Figure S2.**
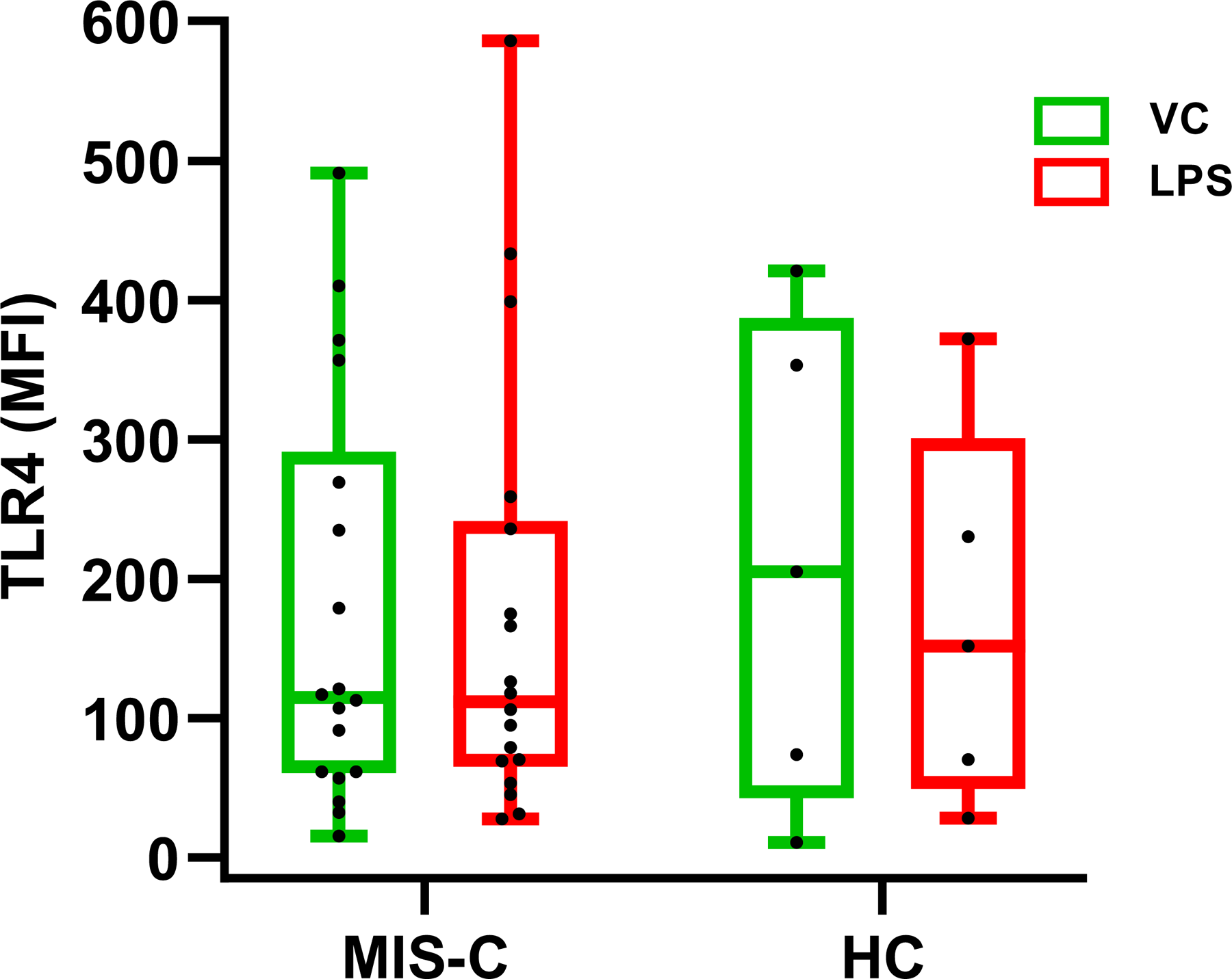
Surface expression of TLR4 on PBMC. PBMC collected from MIS-C patients (n = 17) and healthy controls (HC) (n = 5) were thawed and incubated with lipopolysaccharide (LPS, 50 ng/ml) or endotoxin-free water (vehicle control) for 24 hr, as in Fig. 1. Cells were stained with APC anti-human CD284 (TLR4) antibody as described in *Methods*. Data generated by flow cytometry were expressed as mean fluorescence intensity (MFI). None of the comparisons were found significant (Mann-Whitney U test). MIS-C, MIS-C patients within 30 days of hospital admission; HC, healthy controls.

**Figure S3.**
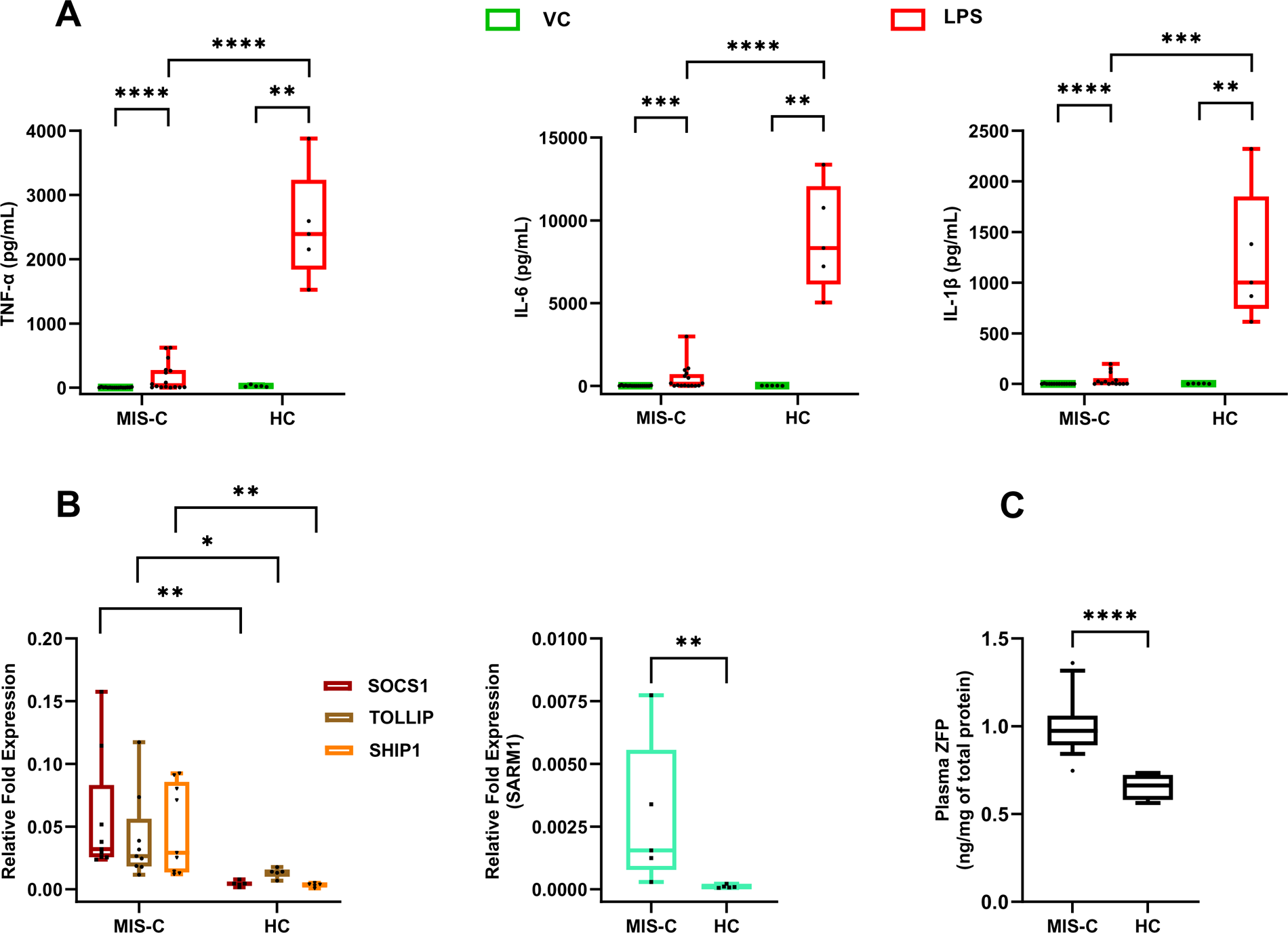
Detection of cellular responses to TLR stimulation and plasma markers. PBMC collected from MIS-C patients >30 days after hospital admission for MIS-C (n = 16) and from healthy controls (HC) (n = 5) were thawed and incubated with lipopolysaccharide (LPS, 50 ng/ml) or endotoxin-free water (vehicle control). Collection of culture supernatants for cytokine release assay and cells for RNA extraction was conducted and data are presented as described in the Fig. 1 legend. All data were generated in technical duplicates. **A.** <u>Cytokine release data</u>. Each panel represent one cytokine, as indicated. **B.** <u>Transcript level data by qPCR</u>. Data are divided in two panels according to relative transcript abundance. **C.** <u>Plasma zonulin levels</u>. The levels of zonulin in plasma were measured, expressed, and presented as in Fig. 2D.

**Figure S4.**
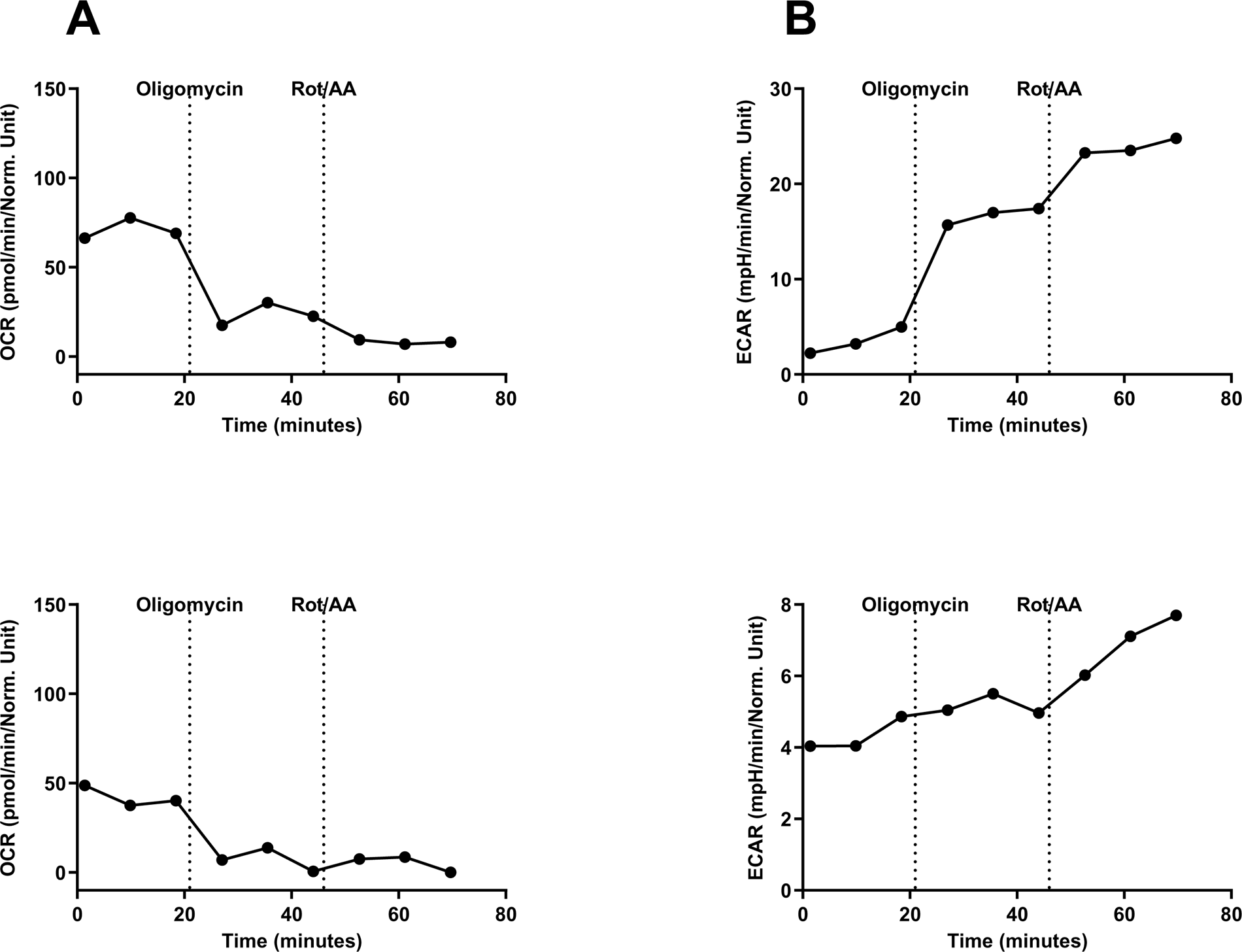
Metabolic Profiles by Seahorse assays. **A.** <u>Oxygen consumption rates</u> (OCR) and **B.** <u>Extracellular acidification rates</u> (ECAR) for a representative healthy control subject (upper panels) and MIS-C subject (lower panels) at baseline and in response to mitochondrial inhibitors.

## Notes

### Competing Interest Statement

The authors have declared no competing interest.

### Summary of Updates

Change in Supplementary Table S4 Change in Figure legend of Figure S4

## References

1. Götzinger F, Santiago-García B, Noguera-Julián A, Lanaspa M, Lancella L, Calò Carducci FI, et al. COVID-19 in children and adolescents in Europe: a multinational, multicentre cohort study. Lancet Child Adolesc Health. 2020;4(9):653–61.

2. Viner RM, Mytton OT, Bonell C, Melendez-Torres GJ, Ward J, Hudson L, et al. Susceptibility to SARS-CoV-2 Infection Among Children and Adolescents Compared With Adults: A Systematic Review and Meta-analysis. JAMA Pediatr. 2021;175(2):143–56.

3. Tsankov BK, Allaire JM, Irvine MA, Lopez AA, Sauvé LJ, Vallance BA, et al. Severe COVID-19 Infection and Pediatric Comorbidities: A Systematic Review and Meta-Analysis. Int J Infect Dis. 2021;103:246–56.

4. Chou J, Thomas PG, and Randolph AG. Immunology of SARS-CoV-2 infection in children. Nat Immunol. 2022;23(2):177–85.

5. Riphagen S, Gomez X, Gonzalez-Martinez C, Wilkinson N, and Theocharis P. Hyperinflammatory shock in children during COVID-19 pandemic. The Lancet. 2020;395(10237):1607–8.

6. Centers for Disease Control and Prevention. Emergency preparedness and response: multisystem inflammatory syndrome in children (MIS-C) associated with coronavirus disease 2019 (COVID-19). CDC Emergency Preparedness and Response https://emergency.cdc.gov/han/2020/han00432.asp (2021).

7. https://www.who.int/publications/i/item/multisystem-inflammatory-syndrome-in-children-and-adolescents-with-covid-19.

8. Brodin P. Immune responses to SARS-CoV-2 infection and vaccination in children. Semin Immunol. 2023;69:101794.

9. Feleszko W, Okarska-Napierala M, Buddingh EP, Bloomfield M, Sediva A, Bautista-Rodriguez C, et al. Pathogenesis, immunology, and immune-targeted management of the multisystem inflammatory syndrome in children (MIS-C) or pediatric inflammatory multisystem syndrome (PIMS): EAACI Position Paper. Pediatr Allergy Immunol. 2023;34(1):e13900.

10. Henderson LA, and Yeung RSM. MIS-C: early lessons from immune profiling. Nat Rev Rheumatol. 2021;17(2):75–6.

11. de Cevins C, Luka M, Smith N, Meynier S, Magerus A, Carbone F, et al. A monocyte/dendritic cell molecular signature of SARS-CoV-2-related multisystem inflammatory syndrome in children with severe myocarditis. Med. 2021;2(9):1072–92 e7.

12. Lee PY, Platt CD, Weeks S, Grace RF, Maher G, Gauthier K, et al. Immune dysregulation and multisystem inflammatory syndrome in children (MIS-C) in individuals with haploinsufficiency of SOCS1. Journal of Allergy and Clinical Immunology. 2020;146(5):1194–200.e1.

13. Chou J, Platt CD, Habiballah S, Nguyen AA, Elkins M, Weeks S, et al. Mechanisms underlying genetic susceptibility to multisystem inflammatory syndrome in children (MIS-C). Journal of Allergy and Clinical Immunology. 2021;148(3):732–8.e1.

14. Lee D, Le Pen J, Yatim A, Dong B, Aquino Y, Ogishi M, et al. Inborn errors of OAS-RNase L in SARS-CoV-2-related multisystem inflammatory syndrome in children. Science. 2023;379(6632):eabo3627.

15. Carter MJ, Fish M, Jennings A, Doores KJ, Wellman P, Seow J, et al. Peripheral immunophenotypes in children with multisystem inflammatory syndrome associated with SARS-CoV-2 infection. Nat Med. 2020.

16. Gruber CN, Patel RS, Trachtman R, Lepow L, Amanat F, Krammer F, et al. Mapping Systemic Inflammation and Antibody Responses in Multisystem Inflammatory Syndrome in Children (MIS-C). Cell. 2020;183(4):982–95.e14.

17. Lee PY, Day-Lewis M, Henderson LA, Friedman KG, Lo J, Roberts JE, et al. Distinct clinical and immunological features of SARS-CoV-2-induced multisystem inflammatory syndrome in children. J Clin Invest. 2020;130(11):5942–50.

18. Sacco K, Castagnoli R, Vakkilainen S, Liu C, Delmonte OM, Oguz C, et al. Immunopathological signatures in multisystem inflammatory syndrome in children and pediatric COVID-19. Nat Med. 2022;28(5):1050–62.

19. Vella LA, Giles JR, Baxter AE, Oldridge DA, Diorio C, Kuri-Cervantes L, et al. Deep immune profiling of MIS-C demonstrates marked but transient immune activation compared to adult and pediatric COVID-19. Sci Immunol. 2021;6(57).

20. Ramaswamy A, Brodsky NN, Sumida TS, Comi M, Asashima H, Hoehn KB, et al. Immune dysregulation and autoreactivity correlate with disease severity in SARS-CoV-2-associated multisystem inflammatory syndrome in children. Immunity. 2021;54(5):1083–95 e7.

21. Consiglio CR, Cotugno N, Sardh F, Pou C, Amodio D, Rodriguez L, et al. The Immunology of Multisystem Inflammatory Syndrome in Children with COVID-19. Cell. 2020.

22. Diorio C, Henrickson SE, Vella LA, McNerney KO, Chase J, Burudpakdee C, et al. Multisystem inflammatory syndrome in children and COVID-19 are distinct presentations of SARS-CoV-2. J Clin Invest. 2020;130(11):5967–75.

23. Kawai T, and Akira S. The role of pattern-recognition receptors in innate immunity: update on Toll-like receptors. Nat Immunol. 2010;11(5):373–84.

24. Kagan JC, and Barton GM. Emerging principles governing signal transduction by pattern-recognition receptors. Cold Spring Harb Perspect Biol. 2014;7(3):a016253.

25. Kawai T, and Akira S. TLR signaling. Semin Immunol. 2007;19(1):24–32.

26. Kawai T, and Akira S. Signaling to NF-kappaB by Toll-like receptors. Trends Mol Med. 2007;13(11):460–9.

27. Yonker LM, Gilboa T, Ogata AF, Senussi Y, Lazarovits R, Boribong BP, et al. Multisystem inflammatory syndrome in children is driven by zonulin-dependent loss of gut mucosal barrier. J Clin Invest. 2021;131(14).

28. Windhorst DB, Zelickson AS, and Good RA. Chediak-Higashi syndrome: hereditary gigantism of cytoplasmic organelles. Science. 1966;151(3706):81–3.

29. Burkhardt JK, Wiebel FA, Hester S, and Argon Y. The giant organelles in beige and Chediak-Higashi fibroblasts are derived from late endosomes and mature lysosomes. J Exp Med. 1993;178(6):1845–56.

30. Talbert ML, Malicdan MCV, and Introne WJ. Chediak-Higashi syndrome. Curr Opin Hematol. 2023;30(4):144–51.

31. Kaplan J, De Domenico I, and Ward DM. Chediak-Higashi syndrome. Curr Opin Hematol. 2008;15(1):22–9.

32. Risma KA, and Marsh RA. Hemophagocytic Lymphohistiocytosis: Clinical Presentations and Diagnosis. J Allergy Clin Immunol Pract. 2019;7(3):824–32.

33. Bloch C, Jais JP, Gil M, Boubaya M, Lepelletier Y, Bader-Meunier B, et al. Severe adult hemophagocytic lymphohistiocytosis (HLHa) correlates with HLH-related gene variants. J Allergy Clin Immunol. 2024;153(1):256–64.

34. Poltorak A, He X, Smirnova I, Liu MY, Van Huffel C, Du X, et al. Defective LPS signaling in C3H/HeJ and C57BL/10ScCr mice: mutations in Tlr4 gene. Science. 1998;282(5396):2085-8.

35. Chanteux H, Guisset AC, Pilette C, and Sibille Y. LPS induces IL-10 production by human alveolar macrophages via MAPKinases- and Sp1-dependent mechanisms. Respir Res. 2007;8(1):71.

36. Chang EY, Guo B, Doyle SE, and Cheng G. Cutting edge: involvement of the type I IFN production and signaling pathway in lipopolysaccharide-induced IL-10 production. J Immunol. 2007;178(11):6705–9.

37. Medvedev AE, Sabroe I, Hasday JD, and Vogel SN. Tolerance to microbial TLR ligands: molecular mechanisms and relevance to disease. J Endotoxin Res. 2006;12(3):133–50.

38. Biswas SK, and Lopez-Collazo E. Endotoxin tolerance: new mechanisms, molecules and clinical significance. Trends Immunol. 2009;30(10):475–87.

39. Piao W, Song C, Chen H, Diaz MA, Wahl LM, Fitzgerald KA, et al. Endotoxin tolerance dysregulates MyD88- and Toll/IL-1R domain-containing adapter inducing IFN-beta-dependent pathways and increases expression of negative regulators of TLR signaling. J Leukoc Biol. 2009;86(4):863–75.

40. Huang S, Liu K, Cheng A, Wang M, Cui M, Huang J, et al. SOCS Proteins Participate in the Regulation of Innate Immune Response Caused by Viruses. Front Immunol. 2020;11:558341.

41. Kwak SG, and Kim JH. Central limit theorem: the cornerstone of modern statistics. Korean J Anesthesiol. 2017;70(2):144–56.

42. Goswami C, Chattopadhyay A, and Chuang EY. Rare variants: data types and analysis strategies. Ann Transl Med. 2021;9(12):961.

43. Rentzsch P, Schubach M, Shendure J, and Kircher M. CADD-Splice-improving genome-wide variant effect prediction using deep learning-derived splice scores. Genome Med. 2021;13(1):31.

44. Szymanski CJ, Humphries WHt, and Payne CK. Single particle tracking as a method to resolve differences in highly colocalized proteins. Analyst. 2011;136(17):3527–33.

45. Wong YC, Kim S, Peng W, and Krainc D. Regulation and Function of Mitochondria-Lysosome Membrane Contact Sites in Cellular Homeostasis. Trends Cell Biol. 2019;29(6):500–13.

46. Mills EL, Kelly B, and O’Neill LAJ. Mitochondria are the powerhouses of immunity. Nat Immunol. 2017;18(5):488–98.

47. Wang Y, Li N, Zhang X, and Horng T. Mitochondrial metabolism regulates macrophage biology. J Biol Chem. 2021;297(1):100904.

48. Feldstein LR, Tenforde MW, Friedman KG, Newhams M, Rose EB, Dapul H, et al. Characteristics and Outcomes of US Children and Adolescents With Multisystem Inflammatory Syndrome in Children (MIS-C) Compared With Severe Acute COVID-19. JAMA. 2021;325(11):1074–87.

49. Song M, Graubard BI, Rabkin CS, and Engels EA. Neutrophil-to-lymphocyte ratio and mortality in the United States general population. Sci Rep. 2021;11(1):464.

50. Khan S, Shafiei MS, Longoria C, Schoggins JW, Savani RC, and Zaki H. SARS-CoV-2 spike protein induces inflammation via TLR2-dependent activation of the NF-kappaB pathway. Elife. 2021;10.

51. Tyrkalska SD, Martinez-Lopez A, Pedoto A, Candel S, Cayuela ML, and Mulero V. The Spike protein of SARS-CoV-2 signals via Tlr2 in zebrafish. Dev Comp Immunol. 2023;140:104626.

52. Zhao Y, Kuang M, Li J, Zhu L, Jia Z, Guo X, et al. SARS-CoV-2 spike protein interacts with and activates TLR41. Cell Res. 2021;31(7):818–20.

53. Fontes-Dantas FL, Fernandes GG, Gutman EG, De Lima EV, Antonio LS, Hammerle MB, et al. SARS-CoV-2 Spike protein induces TLR4-mediated long-term cognitive dysfunction recapitulating post-COVID-19 syndrome in mice. Cell Rep. 2023;42(3):112189.

54. Samsudin F, Raghuvamsi P, Petruk G, Puthia M, Petrlova J, MacAry P, et al. SARS-CoV-2 spike protein as a bacterial lipopolysaccharide delivery system in an overzealous inflammatory cascade. J Mol Cell Biol. 2023;14(9).

55. Vagrecha A, Zhang M, Acharya S, Lozinsky S, Singer A, Levine C, et al. Hemophagocytic Lymphohistiocytosis Gene Variants in Multisystem Inflammatory Syndrome in Children. Biology (Basel). 2022;11(3).

56. Needle RF, and Russell RS. Immunity Debt, a Gap in Learning, or Immune Dysfunction? Viral Immunol. 2023;36(1):1–2.

57. Billard MN, and Bont LJ. Quantifying the RSV immunity debt following COVID-19: a public health matter. Lancet Infect Dis. 2023;23(1):3–5.

58. Chen C, Haupert SR, Zimmermann L, Shi X, Fritsche LG, and Mukherjee B. Global Prevalence of Post-Coronavirus Disease 2019 (COVID-19) Condition or Long COVID: A Meta-Analysis and Systematic Review. J Infect Dis. 2022;226(9):1593–607.

59. Davis HE, McCorkell L, Vogel JM, and Topol EJ. Long COVID: major findings, mechanisms and recommendations. Nat Rev Microbiol. 2023;21(3):133–46.

60. Cheong JG, Ravishankar A, Sharma S, Parkhurst CN, Grassmann SA, Wingert CK, et al. Epigenetic memory of coronavirus infection in innate immune cells and their progenitors. Cell. 2023;186(18):3882–902 e24.

61. Plotegher N, and Duchen MR. Mitochondrial Dysfunction and Neurodegeneration in Lysosomal Storage Disorders. Trends Mol Med. 2017;23(2):116–34.

62. Bonam SR, Wang F, and Muller S. Lysosomes as a therapeutic target. Nat Rev Drug Discov. 2019;18(12):923–48.

63. 63. https://www.cdc.gov/mis/mis-c/hcp_cstecdc/.

64. Harris PA, Taylor R, Thielke R, Payne J, Gonzalez N, and Conde JG. Research electronic data capture (REDCap)--a metadata-driven methodology and workflow process for providing translational research informatics support. J Biomed Inform. 2009;42(2):377–81.

65. Harris PA, Taylor R, Minor BL, Elliott V, Fernandez M, O’Neal L, et al. The REDCap consortium: Building an international community of software platform partners. J Biomed Inform. 2019;95:103208.

66. McKenna A, Hanna M, Banks E, Sivachenko A, Cibulskis K, Kernytsky A, et al. The Genome Analysis Toolkit: a MapReduce framework for analyzing next-generation DNA sequencing data. Genome Res. 2010;20(9):1297–303.

67. Wang K, Li M, and Hakonarson H. ANNOVAR: functional annotation of genetic variants from high-throughput sequencing data. Nucleic Acids Res. 2010;38(16):e164.

