## Supplemental Tables for "A genetically modulated Toll-like-receptor-tolerant phenotype in peripheral blood cells of children with multisystem inflammatory syndrome"

| Table S1 \| Demographic, clinical, and laboratory characteristics of MIS-C subjects | |
|---|---|
| | MIS-C (n = 33) |
| **Age** (years) | |
| 0-11 | 17 (51.52%) |
| 12-17 | 16 (48.48%) |
| **Sex** | |
| Female | 14 (42.42%) |
| Male | 19 (57.58%) |
| **Race** | |
| Non-Hispanic White | 11 (33.33%) |
| Non-Hispanic Black | 8 (24.24%) |
| Non-Hispanic Asian | 4 (12.12%) |
| Hispanic | 10 (30.30%) |
| **Clinical History: COVID-related admission** | |
| Median (IQR) Duration of Admission (days) | 4 (3-7) |
| Secondary super infection | 3 (9.09%) |
| New or increased oxygen support or mechanical ventilation | 10 (31.25%) |
| Vasoactive infusions (including ECMO) | 12 (37.50%) |
| Cardiac complications | 16 (50.00%) |
| Neurologic complications | 3 (9.38%) |
| Gastrointestinal complications | 8 (25.00%) |
| Hematologic/thrombotic complications | 8 (25.00%) |
| Renal complications | 1 (3.12%) |
| Received any treatment | 30 (90.91%) |
| **Blood Panel Measurement** | **Median (IQR)** |
| WBC Count (x10^3/µL) LOW | 7.10 (4.19-9.90) |
| Hemoglobin (g/dL) LOW | 9.60 (8.55-10.60) |
| Platelets (x10^3/µL) LOW | 165 (90.50-236) |
| Absolute lymphocyte count (cells/µL) LOW | 1.06 (0.50-2) |
| Absolute neutrophil count (cells/µL) LOW | 5.20 (2.65-8.70) |
| Neutrophil / Lymphocyte Ratio | 4.33 (2.14-12.43) |
| AST/SGOT (U/L) HIGH | 58 (35-109.50) |
| ALT/SGPT (U/L) HIGH | 45.50 (24.50-116) |
| CRP (mg/dL) HIGH | 16.88 (9.57-20.10) |
| Ferritin (ng/mL) HIGH | 673.50 (428.05-1287.75) |
| LDH (U/L) HIGH | 290.50 (262-383) |
| BNP (pg/mL) HIGH | 1196 (446-4304.50) |
| ESR (mm/hr) HIGH | 83 (30-99) |
| D-dimer (mg/L) HIGH | 2079 (774-4611) |
| **Days to Sample Collection** | |
| 0-30 Days | 17 (51.52%) |
| More than 30 Days | 16 (48.48%) |

IQR: Interquartile range; ECMO: Extracorporeal membrane oxygenation; WBC: White blood cell; AST: Aspartate transferase; ALT: Alanine transaminase; CRP: C-reactive protein; LDH: Lactate dehydrogenase; BNP: B-type Natriuretic Peptide; ESR: Erythrocyte sedimentation rate; LOW: Minimum value during hospitalization; HIGH: Maximum value during hospitalization

| **Table S2 \| Cytokine Levels in Plasma: MIS-C Subjects vs Healthy controls** | | | |
|---|---|---|---|
| **Cytokine** | **Healthy Control (n=5)** | **MIS-C (n=17)** | **P-value** |
| **IL-1β** | 0.03 (0.00-0.03) | 0.41 (0.20-0.80) | 0.002 |
| **IL-6** | 0.01 (0.00-0.01) | 0.25 (0.10-0.78) | 0.003 |
| **IL-10** | 0.04 (0.04-0.09) | 0.61 (0.30-0.80) | 0.002 |
| **IL-12p40** | 0.66 (0.61-1.38) | 1.19 (0.56-3.00) | 0.45 |
| **TNFα** | 0.35 (0.16-0.42) | 1.89 (0.92-4.07) | 0.017 |

Blood of MIS-C subjects was collected within 30 days of hospital admission for MIS-C. Values are represented as Median (IQR). Differences across groups were assessed using a Mann-Whitney U test. In addition to key proinflammatory cytokines (IL-6, IL-1β, TNFα), the levels of IL-10 were also increased, in agreement with a potential pro-inflammatory role of this cytokine during SARS-CoV-2 infection and its association with severe outcomes (PMID: 34234112; PMID: 32501293; PMID: 34234779).

| Table S3 \| Cytokine levels following *ex vivo* stimulation | | | | | | | | | | |
|---|---|---|---|---|---|---|---|---|---|---|
| **Cytokine** | **Treatment** | **All MIS-C (n=33) Median (IQR)** | **Age** | | | | | **Sex** | | |
| | | | **0-5 (n=10)** | **6-11 (n=12)** | **12-17 (n=7)** | **18-21 (n=4)** | **P-value** | **Female (n=14)** | **Male (n=19)** | **P-value** |
| **TNF-α** | **VC** | 1.69 (0.91-3.49) | 1.06 (0.89-1.99) | 1.88 (0.62-5.42) | 1.77 (1.67-3.49) | 5.11 (2.13-10.61) | 0.086 | 1.37 (0.78-3.08) | 1.99 (1.29-3.83) | 0.21 |
| | **LPS** | 14.30 (4.27-80.05) | 18.06 (4.27-49.15) | 9.67 (1.78-364.65) | 32.46 (7.43-59.60) | 144.17 (10.54-446.40) | 0.65 | 7.36 (3.07-276.63) | 20.02 (7.43-80.05) | 0.61 |
| **IL-6** | **VC** | 0.31 (0.17-3.06) | 0.26 (0.20-2.70) | 0.74 (0.13-2.99) | 1.44 (0.18-4.66) | 10.84 (0.20-43.24) | 0.64 | 0.26 (0.20-2.70) | 1.23 (0.14-3.46) | 0.94 |
| | **LPS** | 17.05 (3.78-228.10) | 21.26 (3.78-148.37) | 14.58 (1.96-775.10) | 68.19 (3.68-191.02) | 374.95 (7.54-1860.03) | 0.70 | 5.88 (3.68-742.15) | 25.27 (6.50-191.02) | 0.83 |
| **IL-1β** | **VC** | 0 (0-0.42) | 0.08 (0-0.27) | 0 (0-0.33) | 0 (0-0.88) | 0.61 (0.26-4.30) | 0.29 | 0.19 (0-0.32) | 0 (0-0.53) | 0.50 |
| | **LPS** | 1.49 (0.42-22.43) | 1.52 (1.05-6.86) | 0.81 (0-77.92) | 5.01 (0.82-21.05) | 12.39 (0.47-87.65) | 0.93 | 1.21 (0.21-29.94) | 1.77 (0.42-21.05) | 0.99 |
| **IL12-p40** | **VC** | 0 (0-0) | 0 (0-0) | 0 (0-0) | 0 (0-0) | 0 (0-0) | 0.89 | 0 (0-0) | 0 (0-0) | 0.80 |
| | **LPS** | 0 (0-2.53) | 0 (0-1.32) | 0 (0-9.78) | 0 (0-2.50) | 2.84 (0-9.42) | 0.83 | 0 (0-5.68) | 0 (0-2.50) | 0.63 |
| **IL-10** | **VC** | 0 (0-0.46) | 0 (0-0) | 0.17 (0-0.60) | 0 (0-0.62) | 0.41 (0.18-0.53) | 0.13 | 0.17 (0-0.46) | 0 (0-0.48) | 0.50 |
| | **LPS** | 0 (0-1.83) | 0.20 (0-0.91) | 0 (0-14.95) | 0.52 (0-1.83) | 5.28 (0-19.21) | 0.92 | 0.20 (0-10.55) | 0 (0-1.83) | 0.56 |

| Table S3 \| Cytokine levels following *ex vivo* stimulation (continued) | | | | | | | | | |
|---|---|---|---|---|---|---|---|---|---|
| **Cytokine** | **Treatment** | **Race/ Ethnicity** | | | | | **Days to Sample Collection** | | |
| | | **Non-Hispanic White (n=11)** | **Non-Hispanic Black (n=8)** | **Non-Hispanic Asian (n=4)** | **Hispanic (n=10)** | **P-value** | **0-30 Days (n=17)** | **More than 30 Days (n=16)** | **p-value** |
| **TNF-α** | **VC** | 2.29 (0.89-3.83) | 1.83 (0.83-4.31) | 1.39 (0.94-4.61) | 1.73 (1.29-2.57) | 0.94 | 1.21 (0.78-2.29) | 2.40 (1.62-4.83) | 0.040 |
| | **LPS** | 16.10 (2.04-54.13) | 7.60 (2.41-242.54) | 26.71 (3.67-162.89) | 35.66 (7.43-231.75) | 0.84 | 11.72 (1.53-32.46) | 37.08 (8.24-270.44) | 0.078 |
| **IL-6** | **VC** | 2.70 (0.23-11.31) | 0.81 (0.18-2.75) | 0.85 (0.23-11.45) | 0.18 (0.14-0.25) | 0.16 | 0.20 (0.12-1.23) | 1.94 (0.28-7.42) | 0.013 |
| | **LPS** | 25.27 (4.01-148.37) | 9.31 (1.62-492.40) | 115.94 (3.73-485.13) | 99.38 (3.93-479.83) | 0.90 | 12.11 (0.88-68.19) | 89.31 (5.25-668.89) | 0.11 |
| **IL-1β** | **VC** | 0 (0-0.60) | 0 (0-0.23) | 0.30 (0.25-0.51) | 0 (0-0.16) | 0.20 | 0 (0-0.32) | 0.10 (0-0.56) | 0.53 |
| | **LPS** | 1.55 (0.21-6.86) | 1.20 (0-59.82) | 12.66 (0.94-27.11) | 11.21 (0.42-37.97) | 0.85 | 1.05 (0-5.01) | 4.54 (0.67-31.12) | 0.18 |
| **IL12-p40** | **VC** | 0 (0-0) | 0 (0-0.68) | 0 (0-0) | 0 (0-0) | 0.28 | 0 (0-0) | 0 (0-0) | 0.083 |
| | **LPS** | 0 (0-1.30) | 0 (0-8.29) | 1.26 (0-4.10) | 1.25 (0-4.35) | 0.74 | 0 (0-1.36) | 0.65 (0-6.46) | 0.26 |
| **IL-10** | **VC** | 0 (0-0) | 0.17 (0-0.48) | 0.19 (0-0.49) | 0.42 (0-0.62) | 0.077 | 0 (0-0.36) | 0 (0-0.54) | 0.67 |
| | **LPS** | 0 (0-0.71) | 0 (0-8.59) | 0.66 (0.20-5.73) | 0.91 (0-10.54) | 0.69 | 0 (0-0.55) | 0.36 (0-11.64) | 0.29 |

VC: Vehicle control; LPS: Lipopolysaccharide. Data are presented as medians (IQR). Differences across groups were tested using a Mann-Whitney U test for dichotomous categories and Kruskal-Wallis test of medians for categories with 3 or more groups.

**Table S4 | Target genes related to immune functions**

| | | | | | | | | | | |
|---|---|---|---|---|---|---|---|---|---|---|
| ACD | ACP5 | ACTB | ACTR2 | ADA | ADA2 | ADAM17 | ADAR | ADGRL2 | AGA | AICDA |
| AIRE | AK2 | ALG13 | ANGTP2 | AP1S3 | AP3B1 | AP3D1 | APOL1 | APRIL | ARMC5 | ARPC1B |
| ATG9 | ATM | ATP6AP1 | B2M | BACH2 | BANK1 | BCL6 | BCL10 | BCL11B | BLK | BLM |
| BLNK | BLOC1S6 | BTK | BTN3A1 | BTNL2 | BTNL3 | BTNL8 | C1QA | C1QB | C1QC | C1R |
| C1S | C2 | C3 | C4A | C4B | C5 | C6 | C7 | C8A | C8B | C8G |
| C9 | CARD9 | CARD11 | CARD14 | CARMIL2 | CASP3 | CASP8 | CASP10 | CCBE1 | CD1C | CDED |
| CD3E | CD3G | CD4 | CD8A | CD19 | CD27 | CD28 | CD40 | CD40LG | CD46 | CD55 |
| CD59 | CD70 | CD79A | CD79B | CD80 | CD81 | CD86 | CD101 | CD247 | CD274 | CD300C |
| CDCA7 | CEBPA | CEBPE | CECR1 | CERS5 | CFB | CFD | CFH | CFHR1 | CFHR2 | CFHR3 |
| CFHR4 | CFHR5 | CFI | CFP | CFTR | CHD7 | CIB1 | CIITA | CLCN7 | CLEC4D | CLEC7A |
| CLPB | CNBP | COLEC11 | COPA | CORO1A | CR2 | CREBBP | CSF2RA | CSF2RB | CSF3 | CSF3R |
| CTC1 | CTLA4 | CTNNBL1 | CTSP1 | CTSC | CXCR4 | CYBA | CYBB | DCLRE1B | DCLRE1C | DDR1 |
| DDX58 | DEF6 | DEPTOR | DGKE | DHFR | DHX9 | DHX16 | DHX58 | DKC1 | DNAJC21 | DNASE1L3 |
| DNASE2 | DNMT3B | DOCK2 | DOCK8 | ELANE | ELF1 | ELF4 | EPCAM | EPG5 | ERBB2 | ERBB2IP |
| ERCC2 | ERCC3 | ERCC6L2 | EXOSC9 | EXTL3 | F12 | FAAP24 | FADD | FAM105B | FAM156A | FAS |
| FASLG | FASTK | FAT4 | FBF1 | FBXL19 | FCGR1A | FCGR2A | FCGR2B | FCGR3A | FCGR3B | FCGRT |
| FCN3 | FERMT3 | FNIP1 | FOXN1 | FOXO3 | FOXP3 | FPR1 | FPR2 | FPR3 | G6PC | G6PC3 |
| G6PD | GAD1 | GATA1 | GATA2 | GFI1 | GIMAP5 | GIMAP6 | GIMAP8 | GINS1 | GJC2 | GNAI2 |
| GSDMB | GTF2H4 | GTF2H5 | GTF2I | GUCY2C | H2AFX | HAX1 | HCG14 | HELLS | HMOX1 | HPS1 |
| HPS4 | HPS6 | HRAS | HTRA2 | HYOU1 | IBTK | ICOS | ICOSLG | IFIH1 | IFNAR1 | IFNAR2 |
| IFNGR1 | IFNGR2 | IFNLR1 | IGF1R | IGHG2 | IGHM | IGKC | IGLL1 | IKBKB | IKBKG | IKZF1 |
| IKZF2 | IKZF3 | IKZF4 | IKZF5 | IL1RN | IL2 | IL2RA | IL2RG | IL6R | IL6ST | IL7R |
| IL10 | IL10RA | IL10RB | IL12B | IL12RB1 | IL12RB2 | IL15RA | IL17A | IL17F | IL17RA | IL17RC |
| IL18 | IL18BP | IL19 | IL21 | IL21R | IL22 | IL23A | IL31RA | IL36RN | INO80 | INPP5D |
| INSR | IRAK1 | IRAK4 | IRF1 | IRF2 | IRF2BP2 | IRF3 | IRF4 | IRF5 | IRF6 | IRF7 |
| IRF8 | IRF9 | IRG1 | IRS4 | ISG15 | ITCH | ITGAM | ITGAV | ITGB2 | ITK | JAGN1 |
| JAK1 | JAK2 | JAK3 | KDM6A | KMT2A | KMT2D | KRAS | LACC1 | LAMTOR2 | LAT | LCK |
| LCP2 | LIG1 | LIG4 | LPIN2 | LRBA | LRCH4 | LRR3C | LRRC8A | LYN | LYST | LYZ |
| MAGT1 | MAL | MALT1 | MAN1C1 | MAN2B1 | MAN2B2 | MANBA | MAP3K14 | MAPK12 | MASP1 | MASP2 |
| MBL2 | MC2R | MCM4 | MDC1 | MED24 | MEFV | MR4728 | MIR6884 | MKL1 | MLPH | MOGS |
| MPHOSPH6 | MPI | MPO | MRE11 | MRE11A | MS4A1 | MSH6 | MSN | MST1 | MTHFD1 | MTOR |
| MVK | MYD88 | MYO5A | MYO5B | MYSM1 | N4BP1 | NBAS | NBN | NCF1 | NCF2 | NCF4 |
| NCSTN | NDNL2 | NFAT5 | NFATC1 | NFKB1 | NFKB2 | NFKBIA | NFKBID | NHEJ1 | NHP2 | NKX2-5 |
| NLRC4 | NLRP1 | NLRP3 | NLRP7 | NLRP12 | NOD1 | NOD2 | NOP10 | NRAS | NRM | NSMC3E |
| ORAI1 | ORMDL3 | OSTM1 | OTULIN | PARN | PARP12 | PCCA | PCCB | PDCD1 | PDK1 | PEPD |
| PGAP3 | PGM3 | PHPT1 | PIGA | PIK3CD | PIK3CG | PIK3R1 | PIK3R2 | PIK3R3 | PIK3R4 | PIK3R5 |
| PIK3R6 | PLGC1 | PLGC2 | PLEKHM1 | PLG | PMM2 | PMS2 | PNP | POLA1 | POLE | POLE2 |
| POLE3 | POLE4 | POLR3A | POLR3C | POLR3E | POLR3F | POMP | POU2AF1 | POU5F1 | PPP1R1B | PRF1 |
| PRKCD | PRKCQ | PRKD1 | PRKDC | PRM1 | PRM3 | PRPS1 | PSEN1 | PSENEN | PSMA3 | PSMB4 |
| PSMB8 | PSMB9 | PSMB10 | PSMD3 | PSMG2 | PSORS1C3 | PSTPIP1 | PTEN | PTK2B | PTPN7 | PTPN11 |
| PTPN13 | PTPN22 | PTPRC | PTRF | RAB27A | RAC2 | RAD50 | RAG1 | RAG2 | RANBP2 | RASGRP1 |
| RASGRP2 | RBCK1 | RC3H1 | RECQL4 | RELA | RELB | RET | RFX5 | RFXANK | RFXAP | RHOH |
| RIPK1 | RIPK2 | RLTPR | RMI2 | RMRP | RNASEH2A | RNASEH2B | RNASEH2C | RNF31 | RNF168 | RNU4ATAC |
| RORC | RPSA | RTEL1 | RUNX1 | SAMD3 | SAMD9 | SAMD9L | SAMHD1 | SART3 | SBDS | SBNO2 |
| SEMA3E | SERAC1 | SERPING1 | SGPL1 | SH2D1A | SH3BP2 | SKI | SKIV2L | SLC11A | SLC29A3 | SLC35A1 |
| SLC35C1 | SLC37A4 | SLC39A4 | SLC39A7 | SLC46A1 | SLP76 | SMAD3 | SMAD4 | SMARCAL1 | SMARCD2 | SNHG5 |
| SNORA31 | SNORD50A | SNORD50B | SNX10 | SOCS1 | SOS1 | SP110 | SPI1 | SPINK5 | SPPL2A | SPRED2 |
| SRP54 | STARD3 | STAT1 | STAT2 | STAT3 | STAT4 | STAT5A | STAT5B | STAT6 | STIM1 | STK4 |
| STN1 | STX4 | STX11 | STXBP2 | SWAP70 | SYK | TAP1 | TAP2 | TAPBP | TAZ | TBK1 |
| TBX1 | TBX21 | TCF3 | TCF7 | TCIRG1 | TCN2 | TERC | TERT | TFRC | TGFB2 | TGFB3 |
| TGFBR1 | TGFBR2 | THBD | THRA | TICAM1 | TINF2 | TIRAP | TLR1 | TLR2 | TLR3 | TLR4 |
| TLR5 | TLR6 | TLR7 | TLR8 | TLR9 | TLR10 | TMC6 | TMC8 | TMEM173 | TNFAIP3 | TNFRSF1A |
| TNFRSF1B | TNFRSF4 | TNFRSF9 | TNFRSF11A | TNFRSF13B | TNFRSF13C | TNFSF11 | TNFSF12 | TNFSF13 | TOM1 | TPP2 |
| TRAC | TRAF1 | TRAF2 | TRAF3 | TRAF3IP2 | TRAF4 | TRAF5 | TRAF6 | TRAF7 | TREX1 | TREX2 |
| TRIM27 | TRIM60 | TRNT1 | TTC7A | TTC37 | TYK2 | ULK1 | UNC13D | UNC93B1 | UNC119 | UNG |
| USB1 | USP18 | USP43 | VAV1 | VPS13B | VPS45 | WAS | WDR1 | WIPF1 | WIPF2 | WRAP53 |
| XIAP | XRCC5 | ZAP70 | ZBTB24 | ZNF341 | ZNF646 | ZNF668 | ZPBP2 | | | |

**Table S5 | Genes (other than *LYST*) with ≥4 rare variants in the MIS-C cohort**

| Patient ID | Variants (GRCh37) | Gene | HGVS Consequence | MAF gnomAD 2.1.1 | VEP Annotation | CADD | ClinVar |
|---|---|---|---|---|---|---|---|
| **PCI0687** | 4:126337744_C>T | FAT4 | R2329C | 0.000212 | Missense | 21.3 | VUS/Likely Benign |
| **PCI0713** | 4:126367606_G>T | FAT4 | S2451I | 0.001707 | Missense | 23 | Benign/Likely Benign |
| **PCI0012** | 4:126373530_A>G | FAT4 | K3787E | 3.98E-06 | Missense | 20.7 | . |
| **PCI0671** | 4:126411403_G>T | FAT4 | G4476W | 3.98E-06 | Missense | 28.5 | . |
| **PCI0667** | 4:126411433_G>A | FAT4 | G4486R | 7.95E-06 | Missense | 22.9 | VUS |
| **PCI0669** | 4:126411808_G>A | FAT4 | A4611T | 3.54E-05 | Missense | 23.7 | VUS |
| **PCI0671** | 2:15415841_ GAACAAGTTTGGA>G | NBAS | S1827_V1830del | 1.77E-05 | Inframe deletion | . | . |
| **PCI0686** | 2:15470732_C>T | NBAS | R1446Q | 4.25E-05 | Missense | 26.1 | Likely Benign |
| **PCI0718** | 2:15523394_A>G | NBAS | M1102T | 3.98E-06 | Missense | 25.4 | VUS |
| **PCI0012** | 2:15534391_G>A | NBAS | R1073C | 0.003513 | Missense | 25.1 | Benign/Likely Benign |
| **PCI0590** | 2:15698706_C>T | NBAS | R57Q | 0 | 0.02 | 22.4 | . |
| **PCI0716** | 17:56348108_TTGTTC TTAGACACGGTGGTG ATGCCTGTGTTGTCG CAGATGATCCGGGG CAATGAGATCTGGGC CAGGGCCTGTCGC>T | MPO | R691Hfs*34 | 0 | Frameshift | . | . |
| **PCI0705** | 17:56355353_C>G | MPO | E347Q | 0.000177 | Missense | 20.2 | . |
| **PCI0014** | 17:56357382_G>C | MPO | c.249-7C>G | 1.21E-05 | Splice region | . | . |
| **PCI0012** | 17:56357805_A>C | MPO | V57G | 3.99E-06 | Missense | 31 | . |
| **PCI0014** | 8:48691175_G>C | PRKDC | L3899V | 0.000601 | Missense | 21.2 | VUS/ Likely Benign |
| **PCI0668** | 8:48730007_T>G | PRKDC | c.9554+4A>C | 0.000583 | Splice region | . | Likely Benign |
| **PCI0709** | 8:48752626_A>G | PRKDC | C2468R | 0.000526 | Missense | 23.9 | Likely Benign |
| **PCI0712** | 8:48805870_C>G | PRKDC | G1226R | 1.24E-05 | Missense | 24.8 | VUS |
| **PCI0686** | 11:126162572_G>A | TIRAP | V90M | 3.20E-05 | Missense | 22.9 | . |
| **PCI0594** | 11:126162704_T>C | TIRAP | C134R | 0.000286 | Missense | 25.9 | . |
| **PCI0714** | 11:126162795_C>A | TIRAP | A164D | 3.56E-05 | Missense | 26.6 | . |
| **PCI0714** | 11:126162832_C>G | TIRAP | I176M | 5.32E-05 | Missense | 24 | . |
| **PCI0010** | 8:145738410_T>TGGT GCA | RECQL4 | C857_T858dup | 0.00217 | Inframe insertion | . | Benign/Likely Benign |
| **PCI0711** | 8:145738796_G>A | RECQL4 | Q757* | 0.000118 | Missense | 49 | Pathogenic |
| **PCI0012** | 8:145739416_C>T | RECQL4 | V652M | 0.002460 | Missense | 20.3 | VUS/Benign/ Likely Benign |
| **PCI0708** | 8:145741453_CCT>C | RECQL4 | R350Gfs*21 | 9.31E-05 | Frameshift | . | Pathogenic/ Likely Pathogenic |

Genomic sequencing was performed on MIS-C patient samples. Data were analyzed for coding and splice site variants with a minor allele frequency (MAF) ≤ 0.005 from the gnomAD 2.1.1 database, with additional filtering for a predicted CADD score ≥20 for missense variants, and small indels. VUS = variant of uncertain significance.

| Table S6 \| Rare non-synonymous *LYST* gene variants in the MIS-C cohort | | | | | | | | | | | |
|---|---|---|---|---|---|---|---|---|---|---|---|
| **Patients** | **Variants (GRCh37)** | **HGVS NM_000081** | **VEP Annotation** | **MAF gnomAD v2.1.1** | **CADD** | **SIFT-4G** | **Polyphen2 HDIV** | **Splicing dbscSNV11** | **regSNP -intron** | **ClinVar** | **dbSNP** |
| **PCI0014** | 1:235840850_C>T | c.10870G>A (p.V3624I) | Missense | 1.19E-05 | 26.5 | D | D | . | . | VUS | rs776033238 |
| **PCI0593** | 1:235894366_A>C | c.8913T>G (p.N2971K) | Missense | 2.48E-03 | 22.4 | T | P | . | . | VUS/ Likely benign | rs34702903 |
| **PCI0594** | 1:235940532_C>G | c.5291G>C (p.G1764A) | Missense | 1.31E-03 | 22.8 | D | P | . | . | Benign/ Likely benign | rs35413645 |
| **PCI0683** | 1:235966237_T>C | c.3683A>G (p.N1228S) | Missense | 4.99E-04 | 22 | D | P | . | . | VUS/ Likely benign | rs145553827 |
| | 1:235969077_C>A | c.3359G>T (p.S1120I) | Missense | 1.02E-03 | 14.89 | T | B | . | . | Benign/ Likely benign | rs143223086 |
| **PCI0668** | 1:235993575_T>C | c.143A>G (p.H48R) | Missense | 3.08E-04 | 10.56 | T | B | . | . | VUS | rs200132460 |
| **PCI0705** | 1:235993701_T>C | c.17A>G (p.N6S) | Missense | 4.00E-06 | 24.9 | D | D | . | . | . | rs1254846711 |
| **PCI0589** | 1:235967973_A>C | c.3394-8T>G (p.?) | Splice region | 0 | . | . | . | 0.38 | D | . | . |

Genomic sequencing was performed on MIS-C patient samples. Data were analyzed for coding and splice site variants with a minor allele frequency (MAF) < 0.005. A total of six MIS-C patients had rare missense variants (5 with CADD >20) and one with a predicted splice variant in *LYST*. CADD, SIFT4G, and Polyphen2 HDIV are in silico predictors of variant pathogenicity for missense variants; Splicing dbscSNV11 and regSNP-intron are in silico predictors of splice site effects. For SIFT4G, D = deleterious, T = tolerated. For Polyphen2 HDIV, D = damaging, P = possibly damaging, B = benign. VUS = variant of uncertain significance.

**Table S7 | Common non-synonymous *LYST* gene variants in the MIS-C cohort**

| Patients | Variants (GRCh37) | HGVS NM_000081 | VEP Annotation | MAF gnomAD v2.1.1 | CADD | SIFT-4G | Polyphen2 HDIV | ClinVar | dbSNP |
|---|---|---|---|---|---|---|---|---|---|
| **PCI0667** | 1:235897907_C>T | c.8411G>A (p.G2804D) | missense | 0.013 | 11.29 | T | B | Benign | rs35333195 |
| | 1:235909815_A>T | c.7793T>A (p.F2598Y) | missense | 7.51E-03 | 19.98 | D | P | Benign | rs34642241 |
| | 1:235963637_T>G | c.3989A>C (p.D1330A) | missense | 0.013 | 16.02 | T | B | Benign/ Likely benign | rs74641549 |
| | 1:235969386_C>T | c.3050G>A (p.S1017N) | missense | 0.014 | 0.001 | T | B | Benign | rs10465613 |
| | 1:235973544_A>C | c.574T>G (p.L192V) | missense | 0.014 | 7.079 | T | B | Benign | rs7524261 |
| **PCI0714** | 1:235897907_C>T | c.8411G>A (p.G2804D) | missense | 0.013 | 11.29 | T | B | Benign | rs35333195 |
| | 1:235909815_A>T | c.7793T>A (p.F2598Y) | missense | 7.51E-03 | 19.98 | D | P | Benign | rs34642241 |
| | 1:235963637_T>G | c.3989A>C (p.D1330A) | missense | 0.013 | 16.02 | T | B | Benign/ Likely benign | rs74641549 |
| | 1:235969386_C>T | c.3050G>A (p.S1017N) | missense | 0.014 | 0.001 | T | B | Benign | rs10465613 |
| | 1:235973544_A>C | c.574T>G (p.L192V) | missense | 0.014 | 7.079 | T | B | Benign | rs7524261 |
| **PCI0708** | 1:235897907_C>T | c.8411G>A (p.G2804D) | missense | 0.013 | 11.29 | T | B | Benign | rs35333195 |
| | 1:235963637_T>G | c.3989A>C (p.D1330A) | missense | 0.013 | 16.02 | T | B | Benign/ Likely benign | rs74641549 |
| | 1:235969386_C>T | c.3050G>A (p.S1017N) | missense | 0.014 | 0.001 | T | B | Benign | rs10465613 |
| | 1:235973544_A>C | c.574T>G (p.L192V) | missense | 0.014 | 7.079 | T | B | Benign | rs7524261 |
| **PCI0709** | 1:235897907_C>T | c.8411G>A (p.G2804D) | missense | 0.013 | 11.29 | T | B | Benign | rs35333195 |
| | 1:235963637_T>G | c.3989A>C (p.D1330A) | missense | 0.013 | 16.02 | T | B | Benign/ Likely benign | rs74641549 |
| | 1:235969386_C>T | c.3050G>A (p.S1017N) | missense | 0.014 | 0.001 | T | B | Benign | rs10465613 |
| | 1:235973544_A>C | c.574T>G (p.L192V) | missense | 0.014 | 7.079 | T | B | Benign | rs7524261 |
| **PCI0594** | 1:235933535_C>G | c.5847G>C (p.Q1949H) | missense | 0.019 | 15.77 | T | B | Benign | rs6665568 |
| **PCI0683** | 1:235933535_C>G | c.5847G>C (p.Q1949H) | missense | 0.019 | 15.77 | T | B | Benign | rs6665568 |
| **PCI0686** | 1:235933535_C>G | c.5847G>C (p.Q1949H) | missense | 0.019 | 15.77 | T | B | Benign | rs6665568 |
| **PCI0010** | 1:235933535_C>G | c.5847G>C (p.Q1949H) | missense | 0.019 | 15.77 | T | B | Benign | rs6665568 |

Genomic sequencing was performed on MIS-C patient samples. Data were analyzed for coding and splice site variants with a minor allele frequency (MAF) > 0.005. A total of eight MIS-C patients had six different common variants in *LYST*, with four patients sharing the same four variants and four other patients sharing one variant. CADD, SIFT4G, and Polyphen2 HDIV are *in silico* predictors of variant pathogenicity for missense variants. For SIFT4G, D = deleterious, T = tolerated. For Polyphen2 HDIV, D = damaging, P = possibly damaging, B = benign.

| Table S8 \| Demographic and treatment characteristics of MIS-C patients with *LYST* variants | | | | |
|---|---|---|---|---|
| | **All (N=13)** | **Common (N=6)** | **Rare (N=7)** | **P-value** |
| **Age** (years) | | | | 0.66 |
| 0-11 | 4 (57.14%) | 2 (66.67%) | 2 (50.00%) | |
| 12-17 | 3 (42.86%) | 1 (33.33%) | 2 (50.00%) | |
| **Sex** | | | | 0.42 |
| Female | 3 (23.08%) | 2 (33.33%) | 1 (14.29%) | |
| Male | 10 (76.92%) | 4 (66.67%) | 6 (85.71%) | |
| **Race** | | | | 0.040 |
| Non-Hispanic White | 2 (15.38%) | 0 (0.00%) | 2 (28.57%) | |
| Non-Hispanic Black | 6 (46.15%) | 5 (83.33%) | 1 (14.29%) | |
| Non-Hispanic Asian | 5 (38.46%) | 1 (16.67%) | 4 (57.14%) | |
| Hispanic | 4 (57.14%) | 2 (66.67%) | 2 (50.00%) | |
| **Clinical History: Treatments Received during MIS-C hospitalization** | | | | |
| Received any treatment | 12 (92.31%) | 6 (100.00%) | 6 (85.71%) | 0.34 |
| Remdesivir | 2 (15.38%) | 2 (33.33%) | 0 (0.00%) | 0.097 |
| Anakinra | 2 (15.38%) | 1 (16.67%) | 1 (14.29%) | 0.91 |
| Dexamethasone | 9 (69.23%) | 3 (50.00%) | 6 (85.71%) | 0.16 |
| Methylprednisone | 3 (23.08%) | 2 (33.33%) | 1 (14.29%) | 0.42 |
| Anticoagulants | 1 (7.69%) | 0 (0.00%) | 1 (14.29%) | 0.34 |
| IVIG (Intravenous Ig) | 1 (7.69%) | 1 (16.67%) | 0 (0.00%) | 0.26 |
| Other Antibiotics specific for COVID-19 | 3 (23.08%) | 1 (16.67%) | 2 (28.57%) | 0.61 |
| Other treatments for COVID-19 | 12 (92.31%) | 6 (100.00%) | 6 (85.71%) | 0.34 |

Values shown are frequencies (percentages). Differences across groups were tested using a Fisher's Exact test.

| Table S9 \| Laboratory results of MIS-C patients with *LYST* variants | | | | |
|---|---|---|---|---|
| **Blood panel measurement** | **All (N=13)** | **Common (N=6)** | **Rare (N=7)** | **P-value** |
| WBC Count (x10^3/µL) LOW | 8.70 (5.80-14.40) | 9.45 (6.80-17.80) | 7.70 (3.30-11) | 0.52 |
| Hemoglobin (g/dL) LOW | 9.35 (8.25-10.15) | 9.35 (6.90-9.80) | 9.40 (9.20-10.60) | 0.47 |
| Platelets (x10^3/µL) LOW | 185.50 (96.50-317.50) | 217 (195-465) | 136.50 (85-176) | 0.15 |
| Absolute lymphocyte count (cells/µL) LOW | 1.06 (0.46-2.90) | 2.05 (1.06-3.50) | 0.46 (0.07-0.55) | 0.028 |
| Absolute neutrophil count (cells/µL) LOW | 6.61 (2.50-12.20) | 10 (5.20-14.50) | 6.20 (2.50-7.98) | 0.36 |
| Neutrophil / Lymphocyte Ratio | 5 (2.86-26.52) | 4.33 (2.86-5) | 18.52 (6.19-70.26) | 0.46 |
| AST/SGOT (U/L) HIGH | 62.50 (33.50-136) | 82 (40-189) | 49 (27-68) | 0.34 |
| ALT/SGPT (U/L) HIGH | 44.50 (22-55.50) | 47.50 (16-59) | 44 (28-52) | 0.75 |
| CRP (mg/dL) HIGH | 18.33 (11.10-20.10) | 12.61 (3.66-21.50) | 18.86 (17.94-19.50) | 0.52 |
| Ferritin (ng/mL) HIGH | 589 (407.55-1224.50) | 589 (416.10-1100) | 561.50 (399-1887) | 0.87 |
| LDH (U/L) HIGH | 280 (257-362) | 280 (265-323) | 277.50 (251-362) | 0.72 |
| BNP (pg/mL) HIGH | 3089.50 (792-8013) | 3788 (792-4747) | 2391 (1529-8013) | 0.92 |
| ESR (mm/hr) HIGH | 91 (55-124) | 95 (91-130) | 70 (55-85) | 0.20 |
| D-dimer (mg/L,) HIGH | 4.07 (2.17-6.11) | 3.95 (1.19-4.78) | 4.20 (2.59-7.57) | 0.42 |

WBC: White blood cell; AST: Aspartate transferase; ALT: Alanine transaminase; CRP: C-reactive protein; LDH: Lactate dehydrogenase; BNP: B-type Natriuretic Peptide; ESR: Erythrocyte sedimentation rate; LOW: Minimum value during hospitalization; HIGH: Maximum value during hospitalization. Values are represented as Median (IQR), Differences across groups were tested using a Mann-Whitney U test.

**Table S10 | Primer sequences for q-PCR assay**

| Gene | Forward primer (5′–3′) | Reverse primer (5′–3′) |
|---|---|---|
| **TNF-α** | CCCAGGCAGTCAGATCATCTTC | GCTTGAGGGTTTGCTACAACATG |
| **IL-6** | GAAAGCAGCAAAGAGGCACTG | GAAGCATCCATCTT TTTCAGCC |
| **IL-1β** | ACAGATGAAGTGCTCCTTCCA | GTCGGAGATTCGTAGCTGGAT |
| **NFκB1** | GCAGCACTACTTCTTGACCACC | TCTGCTCCTGAGCATTGACGTC |
| **IL-10** | CTGTGAAAACAAGAGCAAGGC | GAAGCTTCTGTTGGCTCCC |
| **IRAK-M** | GCGGGCAAAGTTAAGACCAT | TCCTCAGGCCTTCATCAGAA |
| **SOCS1** | TAGCACACAACCAGGTGGCA | GCTCTGCTGCTGTGGAGACTG |
| **TOLLIP** | GACCACCGTCAGCACTCAG | GGTCATGCCGTAATTCTTGG |
| **SHIP1** | GCGTGCTGTATCGGAATTGC | CACAG GGTATTGCAGATGGG |
| **SARM1** | TGCTCGAC TCTAACCGCTTG | AGGCGTTTCAGGCTCTGGAT |
